# Optimal Systolic Blood pressure control after thrombectomy in acute ischemic stroke-a systematic review and meta-analysis

**DOI:** 10.1101/2024.04.10.24305642

**Authors:** Baikuntha Panigrahi, Rohit Bhatia, Partha Haldar, Risha Sarkar, Imnameren Longkumer

## Abstract

**Background and Objectives:** Although endovascular thrombectomy(EVT) is the standard of care for acute large vessel occlusions(LVO), optimal systolic blood pressure (SBP) control post procedure has remained elusive. Our study aimed to address the question of whether in adult patients of acute ischemic stroke(AIS) who undergo EVT does an intensive SBP control as compared to a less intensive SBP control/conventional control up to 24 hours post procedure lead to a good functional outcomes defined by modified Rankin score(mRS) of 0-2 at 90days.

**Methods:** This PRISMA guidelines were followed for this review. Databases(PubMed, SCOPUS, EMBASE, Google Scholar and Web of Science) were searched for English language articles using predefined search terms till Sep 15 2023. The inclusion criteria consisted of randomized controlled trials (RCTs) and observational studies(at least total 20patients) analysing intensive vs conventional SBP control in AIS due to LVO post-EVT up to 24hours. Studies without a separate comparison primary outcome data, comparing SBP control in AIS due to LVO treated only with IV thrombolysis (IVT) without EVT, case series and case reports were excluded. The primary outcome was the rate of functional independence defined by mRS 0-2 at 90days. Risk of bias was assessed using the New Castle Ottawa scale(NOS) for observational studies, and the revised Cochrane risk-of-bias 2 (ROB2) tool for RCTs.

**Results:** Twelve studies(n=5439 patients-eight observational and four RCTs) were included in the final analysis. The primary outcome was not significantly different between both the arms(RR:1.16;95%CI-0.98-1.37;p=0.08).There were no significant differences in the mortality at 90days(RR:0.83;95%CI-0.68-1.02;p=0.08) and the risk of symptomatic intracranial haemorrhage(RR:0.84;95%CI-0.61-1.16;p=0.29). Fewer patients required decompressive surgery in the intensive arm(RR-0.40; 95%CI-0.25-0.66; p=0.0003). A separate analysis for the primary outcome from pooled observational data favoured intensive control(RR-1.34;95%CI-1.20-1.48; p<0.00001) and data from RCTs favoured conventional control(RR-0.82;95%CI-0.72-0.93;p=0.003).

**Conclusion:** Neither intensive nor conventional SBP control resulted in better functional outcome in the combined analysis of all studies. Although, observational studies favoured intensive control, data from RCTs suggested conventional management as the preferred approach which could currently be a pragmatic strategy. Further ongoing RCTs using homogenous SBP cut-offs will provide more clarity on the ideal SBP target after EVT.

## Introduction

Endovascular thrombectomy with or without thrombolysis is currently the standard of care in patients with acute ischemic stroke due to a large vessel occlusion.^1^ However, even after successful radiological evidence of reperfusion a proportion of patients do not achieve the desired level of functional independence.^2^ This is dependent on numerous modifiable, non-modifiable and unknown factors. Among these, blood pressure (BP) is a major modifiable factor which influences the outcomes after a successful reperfusion.^2,3^ The interaction between BP and outcomes is extremely complex as other critical factors including the collateral status, size of the penumbra and degree of recanalization play a role in the final outcome.^4^ However, the optimal blood pressure after thrombectomy is not clear. The 2019 American Heart Association/American Stroke Association(AHA/ASA) guidelines for acute ischemic stroke recommend that it is reasonable to maintain BP at ≤180/105 mm Hg during and for 24 hours after the procedure (LOE-B-NR;COR-IIa)^1^. It is well acknowledged in the guideline that the data on optimal blood pressure target is sparse and the data supporting this guideline was drawn from blood pressure management protocols in the thrombectomy trials. Since a majority of the patients enrolled in the six hour time window trials had already received thrombolysis, the post thrombolysis blood pressure targets applied to them. Additional data was obtained from the DAWN and ESCAPE trial protocols.^5,6^ The DAWN protocol recommended the systolic BP (SBP) to be maintained at <140mmHg in the first 24hrs after the procedure.^6^ However, the ESCAPE protocol recommended the SBP to be maintained at >150mmHg to maintain collateral perfusion.^5^

Since the last recommendations, multiple randomised and observational data has become available with conflicting results which makes it imperative to perform a systematic review and meta-analysis to carefully address this important question. We conducted this meta-analysis to address the question of whether in adult patients with an acute ischemic stroke due to large vessel occlusion who undergo thrombectomy does an intensive blood pressure control (Target SBP cutoff as defined by the respective study or an upper limit of SBP target for defining intensive control as <140mmHg) as compared to a less intensive blood pressure control/conventional control(higher SBP targets or guideline recommended SBP control) up to 24 hours after the procedure lead to a good functional outcome as defined by modified Rankin score (mRS) of 0-2 at 90 days.

## Methods

### Search strategy

We used the standard PRISMA (Preferred Reporting Items for Systematic Reviews and Meta-Analyses) guidelines for conducting and reporting our review (Checklist in *Supplementary material-1*). The protocol was registered on PROSPERO (https://www.crd.york.ac.uk/prospero/#myprospero) vide registration number-CRD42023463173. An amendment was submitted on 28 January 2024 to define blood pressure targets as per the respective study definition. A comprehensive preliminary search was carried out by three authors (B.P, R.S and I.L) for English Language articles from inception until September 15, 2023, using databases which included PubMed, SCOPUS, EMBASE, Google Scholar, Web of Science and hand searching. The MeSH terms included for the search string were – “Acute stroke”, OR “Ischemic stroke” OR “cerebrovascular accident” OR “cerebral infraction” AND “thrombectomy” OR “endovascular therapy” OR “mechanical aspiration” OR “endovascular technique” OR “endovascular procedure” OR “stent retriever” AND “blood pressure” OR “blood pressure monitor” OR “high blood pressure” OR “low blood pressure” OR “hypertension”. The search results obtained from the various sources were imported to the Covidence software (https://www.covidence.org). Duplicates were removed using the software automatically.

### Screening and Eligibility criteria

Titles and abstracts were reviewed independently by four authors (B.P, RB, R.S, I.L) to evaluate for inclusion and screening of full text. The review question and the PICO statements are summarised in the form of a table (available as *Supplementary material-2*). The inclusion criteria of the studies were-Randomized Controlled Trials (RCTs), observational prospective studies, observational retrospective studies with at least a total of 20 patients, patients included in the study must have had a baseline mRS-0-1,patients with Acute Ischemic Stroke (AIS) due to large vessel occlusion (LVO) involving either the anterior or the posterior circulation and treated with thrombectomy using any technique with or without bridging thrombolytic agent, systemic blood pressure monitoring during and at least 24 hours after the procedure and functional outcomes using the mRS at 90days should have been reported. Intensive control was defined as aggressive control of SBP to a predefined target of <140mmHg or as described in the respective study with the use of anti-hypertensive drugs for up to 24 hours after a successful thrombectomy irrespective of whether the patient received thrombolysis or not. Less Intensive or Conventional blood pressure control was defined as a liberal SBP control strategy where SBP was not lowered until it was >180mmHg as per guidelines. We excluded case series, case reports, studies without a separate comparison outcome data between intensive and less intensive blood pressure groups, studies comparing blood pressure control in AIS due to LVO treated only with Intravenous thrombolysis (IVT) without Endovascular therapy (EVT) and studies without a primary efficacy outcome data. The primary efficacy outcome was the proportion of patients who had a mRS of 0-2 at 90days. Other safety outcomes that were analysed were death at 90 days, development of symptomatic intracranial haemorrhage(sICH) and requirement of decompressive surgery. Any discrepancies were resolved by R.B. Full texts were retrieved for further consideration for inclusion in the study. Individual patient data was not available for the study.

### Data Extraction

The data was extracted on MS Excel (Ver 16.66.1).This included: the name of the journal, year of publication, 1st author’s name, study type-retrospective, prospective study or randomized trial (monocentric or multicentric), number of patients, proportion of patients with a history of hypertension, median NIHSS, stroke localization, intravenous thrombolysis use (if provided), recanalization rates, procedure time, number of patients in the intensive and conventional treatment arm as per the respective study cut-offs, BP parameters measured at least for 24 hours post procedure (mean, minimum, maximum according to the study), rates of symptomatic intracranial haemorrhage (ICH) at 24-36 hours as per the respective study definition and the 3-month mRS according to peri-procedural BP.

### Risk of Bias Assessment

The Newcastle Ottawa Scale (NOS) was used by B.P (reviewed by P.H) to assess the risk of bias in the observational studies. The Risk of Bias (ROB2) tool was used by B.P (reviewed by P.H) for assessing risk of bias in randomized controlled trials.

### Strategy for Data Analysis

Statistical analyses were performed using Review Manager 5.4.1 software by B.P and reviewed by P.H. Random-effects model was used in view of expected heterogeneity to be present due to known differences in stroke patient characteristics, SBP cut-offs and different types of studies(observational and RCTs). The overall effect estimate, and 95% CI was used to generate Forest plots. Heterogeneity was assessed by the Higgin I^2^ statistics. The primary and all secondary efficacy and safety outcomes were assessed by risk ratios (RR). Sensitivity analysis using the leave one study out analysis was done. GRADE pro GDT was used for certainty of evidence assessment.

## Results

### Study selection and evaluation

An initial database search using the MeSH terms yielded 9883 records which included 8958 from PubMed, 1045 from Google Scholar, 545 from Scopus, 288 from EMBASE and 129 from Web of Science. 1026 duplicates were removed(Figure-1). 8857 studies entered the title and abstract screening of which 8713 studies were found to be irrelevant. 72 studies were assessed for full text eligibility and 60 studies were excluded(reasons in the PRISMA flowchart-Figure 1). A total of twelve studies were included in the meta-analysis.

**Figure-1.**
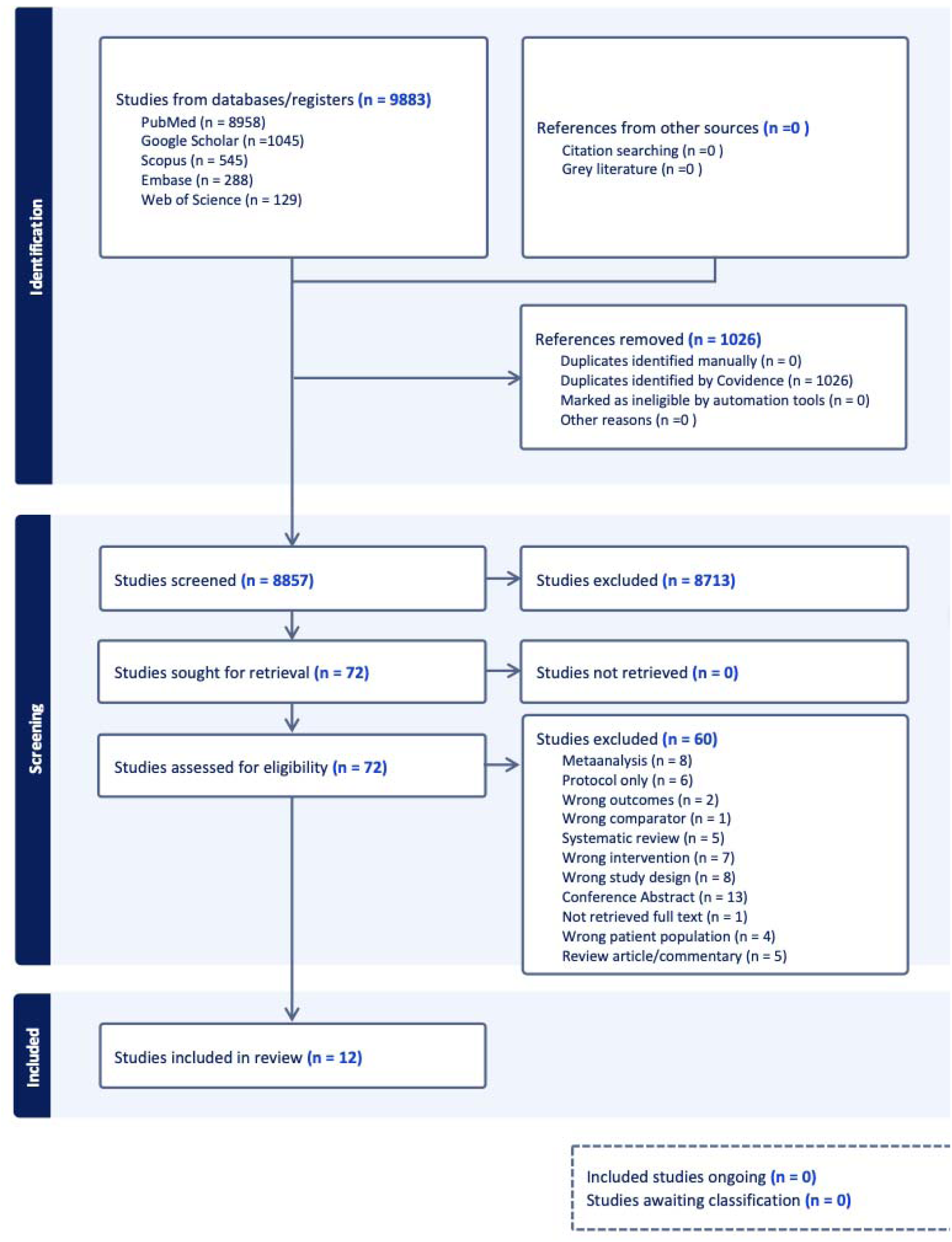
PRISMA flowchart of the study.

### Study Characteristics

Of the twelve studies involving 5439 patients included in the final analysis, eight were observational and four were RCTs.^7–18^ Observational study data for 3868 patients and data from RCTs involving total of 1571 patients was also extracted separately. We used the published data only for conducting the analysis. We did not have access to individual patient data in any of the studies included in the meta-analysis. Five out of the eight observational studies used a SBP cut-off of 140 mmHg to define intensive control.^11,13,15–17^ Two(Chang et al-2019 and P.Upadhyaya et al-2023) studies used a cut-off of 130 mmHg and one study(Eva.A.Mistry et al) used a cut-off of 158mmHg.^12,14,18^ In case of the RCTs the ENCHANTED2/MT used a cut-off SBP of 120mmHg to define its intensive arm.^8^ The BP-TARGET on the other hand used a range of SBP between 100-129mmHg to define its intensive arm.^7^ There were 3 SBP groups in the BEST-II(140,160,180).^9^ For combining the effects we clubbed the <160mmHg and <180mmHg groups into the less intensive group and the <140mmHg as the intensive arm. The OPTIMAL-BP used a SBP cut-off of 140mmHg.^10^ The baseline characteristics and attributes of each study included in this meta-analysis are summarised in the Table-1 and Table-2.

**Table-1.**
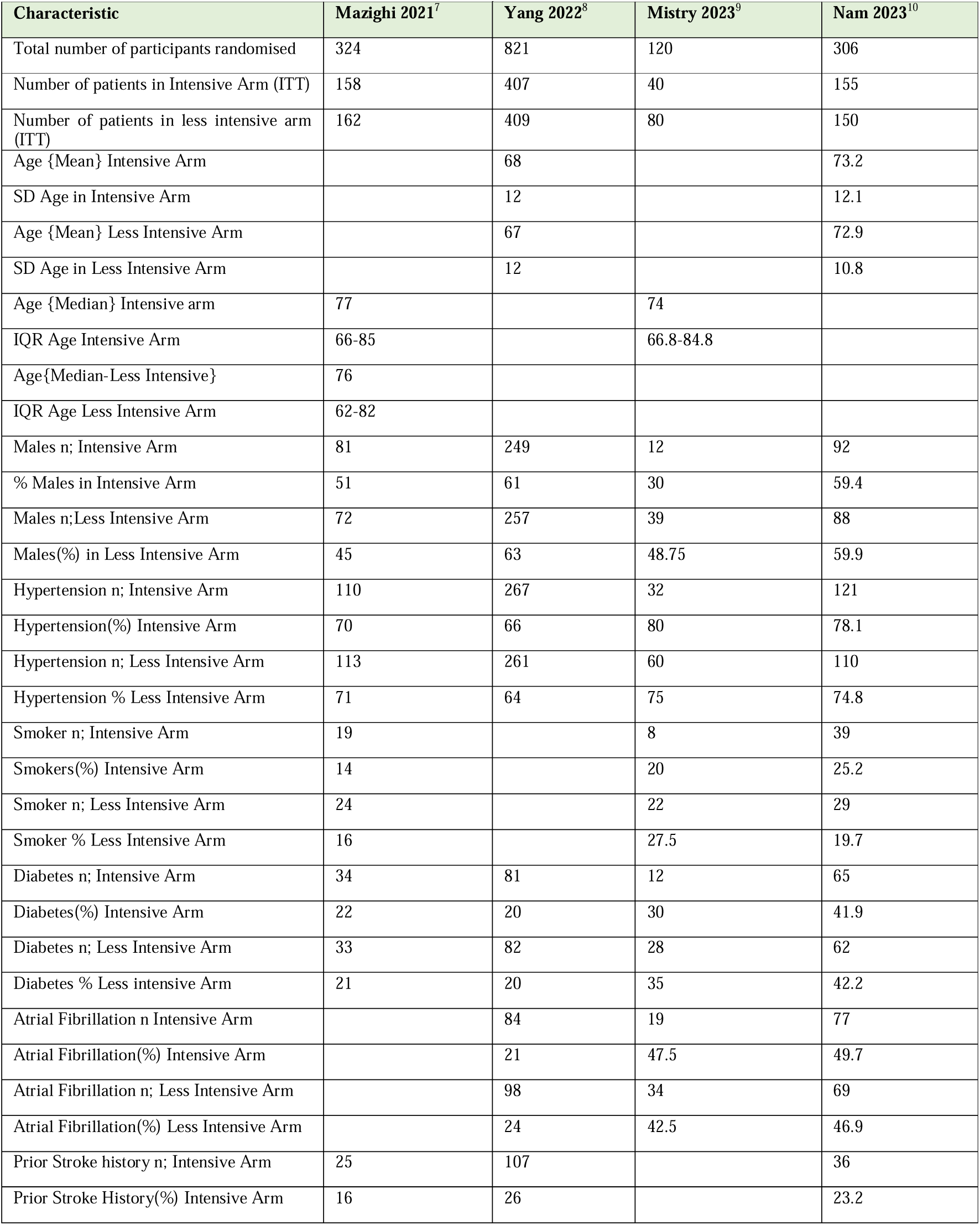
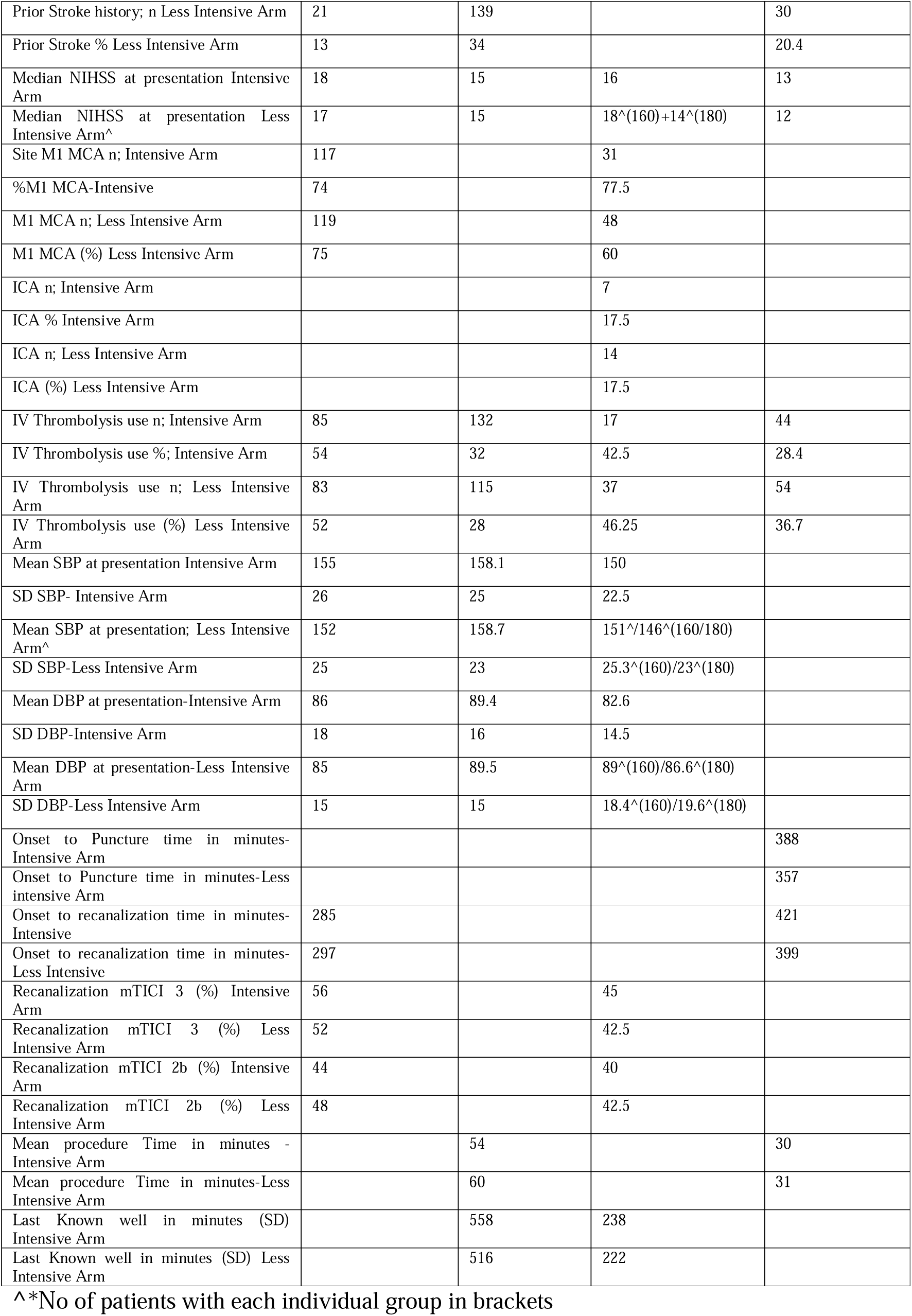
Baseline characteristics of the study population of RCTs.

**Table-2.** Baseline characteristics of the study population of Observational Studies.

| Characteristic | Goyal et al<sup>11</sup> | Eva Mistry et al<sup>12</sup> | Anadani et al<sup>13</sup> | Chang et al<sup>14</sup> | Cernik D et al<sup>15</sup> | Anadani et al<sup>16</sup> | Han et al<sup>17</sup> | P. Upadhyaya et al<sup>18</sup> |
| --- | --- | --- | --- | --- | --- | --- | --- | --- |
| Year | 2017 | 2019 | 2019 | 2019 | 2019 | 2020 | 2021 | 2023 |
| BP Cut-offs | 140 | 158 | 140 | 130 | 140 | 140 | 140 | 130 |
| No of patients-total | 217 | 485 | 1245 | 90 | 690 | 1019 | 187 | 103 |
| n-Intensive | 10 | 195 | 942 | 48 | 376 | 540 | 96 | 44 |
| n-Less intensive | 130 | 290 | 259 | 42 | 314 | 479 | 83 | 59 |
| Age(Mean)-Total | 62 |  | 69 | 72.3 | 71 |  | 60 |  |
| Age(SD)-Total | 14 |  | 14 | 11.8 | 13 |  | 10 |  |
| Age(Mean)_intensive | 61.2 ± 14.2 |  |  | 70.1 |  | 70 |  |  |
| Age(Mean)-Less Intensive |  |  |  | 74.9 |  |  |  |  |
| Age(Median) |  | 69 |  |  |  |  |  | 66 |
| Age(IQR) |  | 57-79 |  |  |  |  |  | 57-75 |
| Males(n) | 108 | 236 | 614 | 54 | 352 | 508 | 157 | 40 |
| Males(%) | 50 | 48.66 | 49.3 | 60 | 51 | 49.8 | 84 | 38.8 |
| Males(Intensive)-n | 2 |  |  | 31 |  | 259 |  | 18 |
| % males-Intensive | 20 |  |  | 64.6 |  | 47.96 |  | 40.9 |
| Males(Less Intensive)-n | 68 |  |  | 23 |  | 249 |  | 22 |
| %males(Less Intensive) | 52.3 |  |  | 54.8 |  | 51.98 |  | 37.3 |
| Admission NIHSS-Baseline-Total | 16(12-21) | 16(11-20) | 16(IQR-7) | 14.7±5.5 | 17 |  | 22(10-34) | 19 |
| Admission NIHSS-Intensive | 17 | 15 |  | 14.2±5.3 |  | 15 |  | 20 |
| Admission NIHSS-Less Intensive* |  | 17 |  | 15.3±5.8 |  | 17*(160)<br>16*(180) |  | 19 |
| Hypertension-Total-n | 166 | 364 | 797 | 43 | 525 | 644 | 133 | 63 |
| Hypertension%-Total | 77 | 75 | 64 | 49.4 | 76 | 62.59 | 71.1 | 61 |
| Hypertension-Intensive(n) | 6 | 122 |  | 20 |  | 326 |  | 29 |
| Hypertension(%)-Intensive | 60 | 62.5 |  | 43.5 |  | 60 |  | 66 |
| Hypertension-Less Intensive-n | 100 | 242 |  | 23 |  | 318 |  | 34 |
| Hypertension(%)-Less Intensive | 76.9 | 83.4 |  | 56.1 |  | 66.4 |  | 59 |
| Diabetes(Total)-n | 70 | 139 | 406 | 25 | 188 | 356 | 51 | 17 |
| Diabetes(Total)-% | 32 | 29 | 33 | 28.7 | 27 | 34.94 | 27.3 | 17 |
| Diabetes(Intensive)-n | 3 | 51 |  | 12 |  | 192 |  | 7 |
| Diabetes(Intensive)% | 30 | 26 |  | 26.1 |  | 36 |  | 16 |
| Diabetes(Less Intensive)-n | 44 | 88 |  | 13 |  | 164 |  | 10 |
| Diabetes(Less Intensive%) | 33.8 | 30 |  | 31.7 |  | 34.2 |  | 18 |
| Afib-Total-n | 54 | 176 | 462 | 51 | 299 |  |  | 31 |
| Afib-Total(%) | 25 | 36 | 37 | 58 | 43 |  |  | 30 |
| Afib(Intensive)-n | 4 | 63 |  | 29 |  | 186 |  | 14 |
| Afib(Intensive(%)) | 40 | 32 |  | 61.7 |  | 34.4 |  | 33 |
| Afib(Less Intensive)-n | 29 | 113 |  | 22 |  | 203 |  | 17 |
| Afib(Less Intensive(%)) | 22.3 | 39 |  | 53.7 |  | 42.4 |  | 30 |
| Prior Stroke-Total(n) |  |  |  |  |  |  | 37 | 13 |
| Prior Stroke-Total% |  |  |  |  |  |  | 19.8 | 13 |
| Prior Stroke-Intensive(n) |  |  |  |  |  |  |  | 6 |
| Prior Stroke-Intensive% |  |  |  |  |  |  |  | 14 |
| Prior Stroke-Less Intensive(n) |  |  |  |  |  |  |  | 7 |
| Prior Stroke-Less Intensive% |  |  |  |  |  |  |  | 12 |
| IVtPA-Total(n) | 141 | 231 | 590 | 54 | 519 | 566 | 36 | 62 |
| IVtPA-Total(%) | 65 | 48 | 55 | 60 | 75 | 55.5 | 19.3 | 60 |
| IVtPA-Intensive(n) | 9 | 107 |  | 29 |  | 278 |  | 27 |
| IVtPA-Intensive(%) | 90 | 55 |  | 60.4 |  | 51 |  | 61 |
| IVtPA-Less Intensive | 84 | 124 |  | 25 |  | 288 |  | 35 |
| IV tPA-Less intensive% | 64.6 | 42 |  | 59.5 |  | 60 |  | 59 |
| ICA-Total(n) | 33 | 129 | 172 | 26 | 160 | 150 |  | 16 |
| ICA-Total(%) | 15 | 27 | 14.2 | 28.9 | 23 | 14.7 |  | 16 |
| ICA-Intensive(n) |  | 49 |  | 11 |  | 66 |  | 8 |
| ICA-Intensive% |  | 25 |  | 22.9 |  | 11 |  | 18 |
| ICA-Less Intensive-n |  | 80 |  | 15 |  | 84 |  | 8 |
| ICA-Less Intensive(%) |  | 27 |  | 35.7 |  | 17.5 |  | 14 |
| MCA-M1-Total(n) | 104 | 309 | 714 | 64 | 567 | 564 |  | 42 |
| MCA-M1-Total(%) | 48 | 64 | 57 | 71.1 | 82 | 55.35 |  | 41 |
| M1-Intensive-n |  | 108 |  | 37 |  | 315 |  | 19 |
| M1 Intensive% |  | 55.3 |  | 77.1 |  | 59 |  | 43 |
| M1 Less Intensive(n) |  | 148 |  | 27 |  | 249 |  | 23 |
| M1 Less Intensive(%) |  | 51 |  | 64.3 |  | 51.98 |  | 39 |
| Post Circulation-Total-n | 38 |  |  |  |  |  | 187 |  |
| Post Circulation-% | 18 |  |  |  |  |  | 100 |  |

\*No of patients with each individual group in brackets

### Risk of Bias

- Observational Studies The Newcastle Ottawa Scale results are summarised in the *Supplementary material-3*. We found all studies to be of good methodological quality with a median of 8 stars.
- RCTs The ROB2 tool demonstrated a “high risk” of bias in the deviations from the intended intervention domain (Domain-2) in three out of four included RCTs. One study had “some concerns” in this domain. The other domains examined had a low risk of bias. The detailed analysis is provided in the *Supplement (Supplementary material-3)*.

### Outcomes

#### Primary Outcome mRS 0-2

Pooled data of all the twelve studies involving 5398 patients (reported outcomes) with 2983 patients in the intensive blood pressure control group and 2415 patients in the conventional control group demonstrated that the primary outcome of mRS-0-2 was seen in 1508/2983(50.55%) patients in the intensive arm as compared to 1106/2415(45.8%) in the conventional arm and was not significantly different between both the arms(RR-1.16; 95%CI-0.98-1.37; p=0.08) as seen in Figure-2.^7–18^

**Figure-2.**
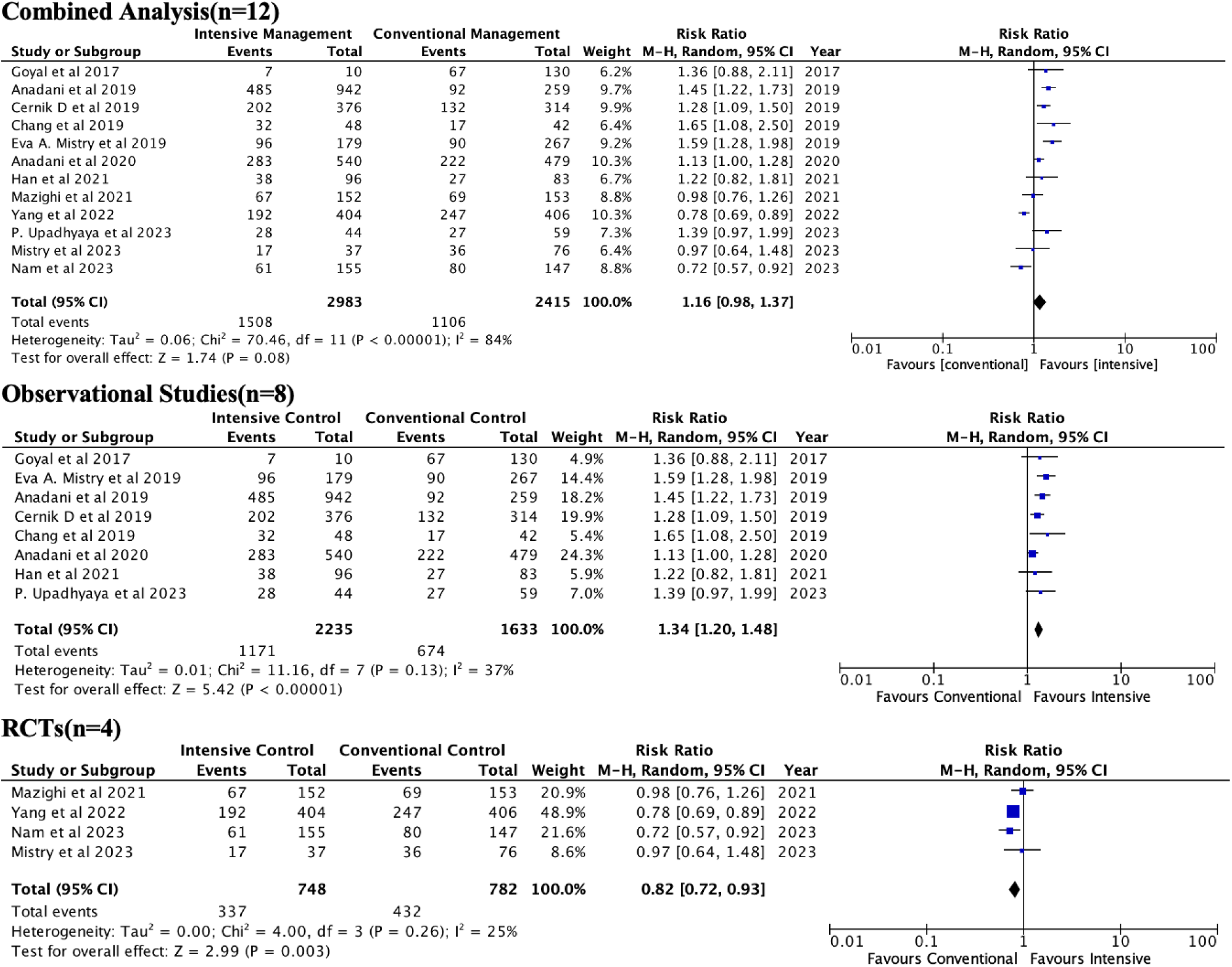
Forest plot for the primary outcome of mRS 0-2 at 90 days. The estimated risk ratio-RR is represented by a square and the corresponding 95% CI are shown for the primary outcome(mRS-0-2) at 90 days. The square’s size is proportional to the study’s weight in the meta-analysis implying studies with larger squares have greater weight. The diamond beneath is the overall pooled estimate and its width denotes the 95%CI around the combined effect size.

Separate analysis of data from eight observational studies showed that mRS 0-2 was seen in 1171 out of 2235 in the intensive control group (52.39%) as compared to 674 out of 1633 patients in the conventional group (41.23%) favouring intensive blood pressure control for a good outcome(RR-1.34; 95%CI-1.20-1.48; p<0.00001) as shown in Figure-2.^11–17^

Similarly, a separate analysis of data from four RCTs showed that the primary outcome was seen in 337/748(45.05%) patients in the intensive arm as compared to 432/782(55.24%) in the conventional arm thus favouring the conventional arm for a good outcome(RR-0.82; 95%CI-0.72-0.93; p=0.003) as shown in Figure-2.^7–10^

#### Death at 90 days

Outcomes for death at 90days were available for 2985 patients in the intensive arm and 2417 patients in the conventional arm from twelve studies. A total of 487/2985(16.3%) patients died in the intensive arm as compared to 485/2417(20.06%) in the conventional arm and the difference between both the arms was not significant(RR-0.83; 95%CI-0.68-1.02; p=0.08) as shown in Figure-3.^7–18^

**Figure-3.**
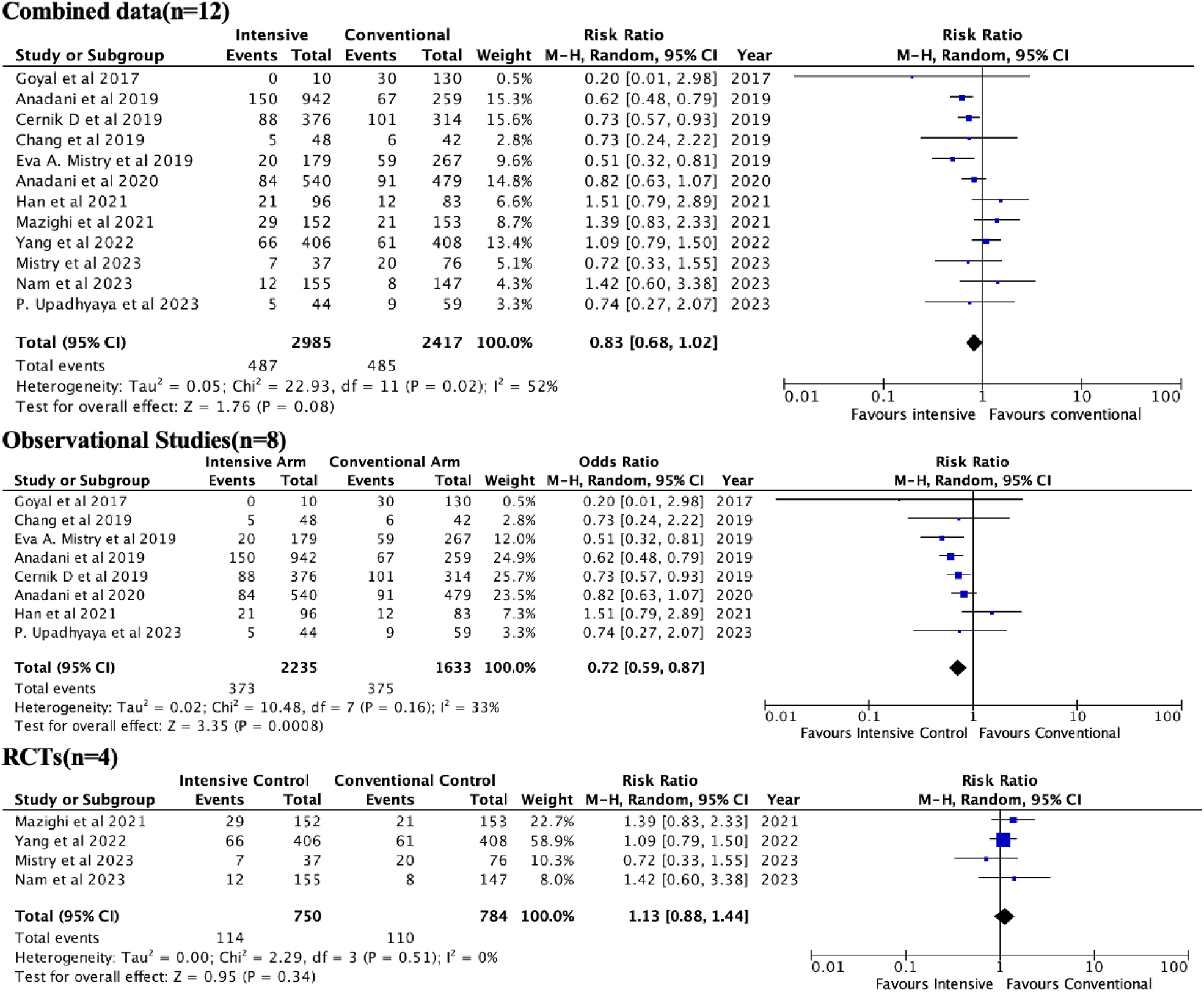
Forest plot for the outcome of death at 90 days. The estimated risk ratio-RR is represented by a square and the corresponding 95% CI are shown for the outcome death at 90 days. The square’s size is proportional to the study’s weight in the meta-analysis implying studies with larger squares have greater weight. The diamond beneath is the overall pooled estimate and its width denotes the 95%CI around the combined effect size.

A separate analysis of data from eight observational studies showed that 373/2235(15.97%) patients died in the intensive arm as compared to 375/1633(22.96%) in the conventional arm. The difference between both the arms was significant with lower risk of dying in the intensive arm(RR-0.72; 95%CI-0.59-0.87; p=0.0008) as shown in Figure-3.^11–17^.

Data from the four RCTs showed that 114/750(15.2%) patients died in the intensive arm as compared to 110/784(14.03%) patients in the conventional arm and the difference between both the arms was not statistically significant (RR-1.13; 95%CI-0.88-1.44; p=0.34) as shown in Figure-3.^7–10^

#### Symptomatic ICH

Data on symptomatic ICH(sICH) was available for nine out of twelve studies involving 2385 patients in the intensive arm and 1797 patients in the conventional arm. A total of 130/2385(5.45%) patients had sICH(as per respective study definition) in the intensive arm compared to 127/1797(7.07%) in the conventional arm and the difference between the two groups was not statistically significant(RR-0.84; 95%CI-0.61-1.16; p=0.29) as shown in Figure-4.^7–11,13,16–18^

**Figure-4.**
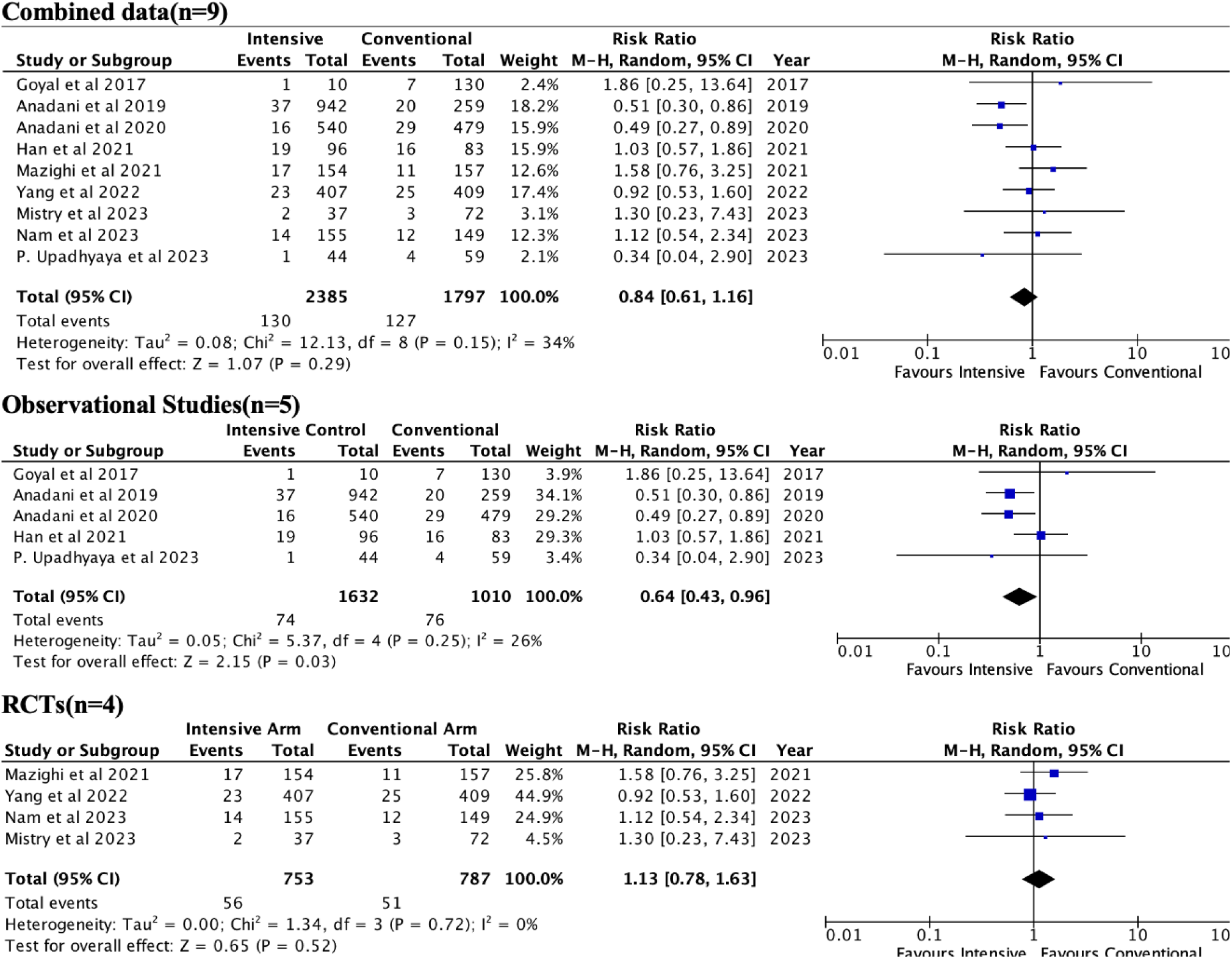
Forest plot for the outcome of symptomatic ICH(sICH) The estimated risk ratio-RR is represented by a square and the corresponding 95% CI are shown for the outcome sICH. The square’s size is proportional to the study’s weight in the meta-analysis implying studies with larger squares have greater weight. The diamond beneath is the overall pooled estimate and its width denotes the 95%CI around the combined effect size.

A separate analysis of data from five observational studies showed that sICH occurred in 74/1632(4.53%) patients in the intensive arm compared to 76/1010(7.52%) patients in the conventional arm and the risk of sICH was significantly lower in the intensive arm compared to the conventional arm(RR-0.64; 95%CI-0.43-0.96; p=0.03) as shown in Figure-4.^11,13,16–18^ Data from the four RCTs showed that symptomatic ICH occurred in 56/753(7.44%) patients in the intensive arm compared to 51/787(6.48%) patients in the conventional arm with no significant differences between the two groups(RR-1.13; 95%CI-0.78-1.63; p=0.52) as shown in Figure-4.^7–10^

### Decompressive surgery

Data on decompressive surgery was available from four out of twelve studies for 1684 patients in the intensive arm as compared to 959 patients in the conventional arm. A total of 44/1684(2.61%) patients in the intensive arm required decompression as compared to 54/959(5.63%) patients in the conventional arm. There was significantly higher requirement of decompressive surgery in the conventional management arm compared to the intensive arm (RR-0.40; 95%CI-0.25-0.66; p=0.0003) as shown in Figure-5.^7,13,16,18^

**Figure-5.**
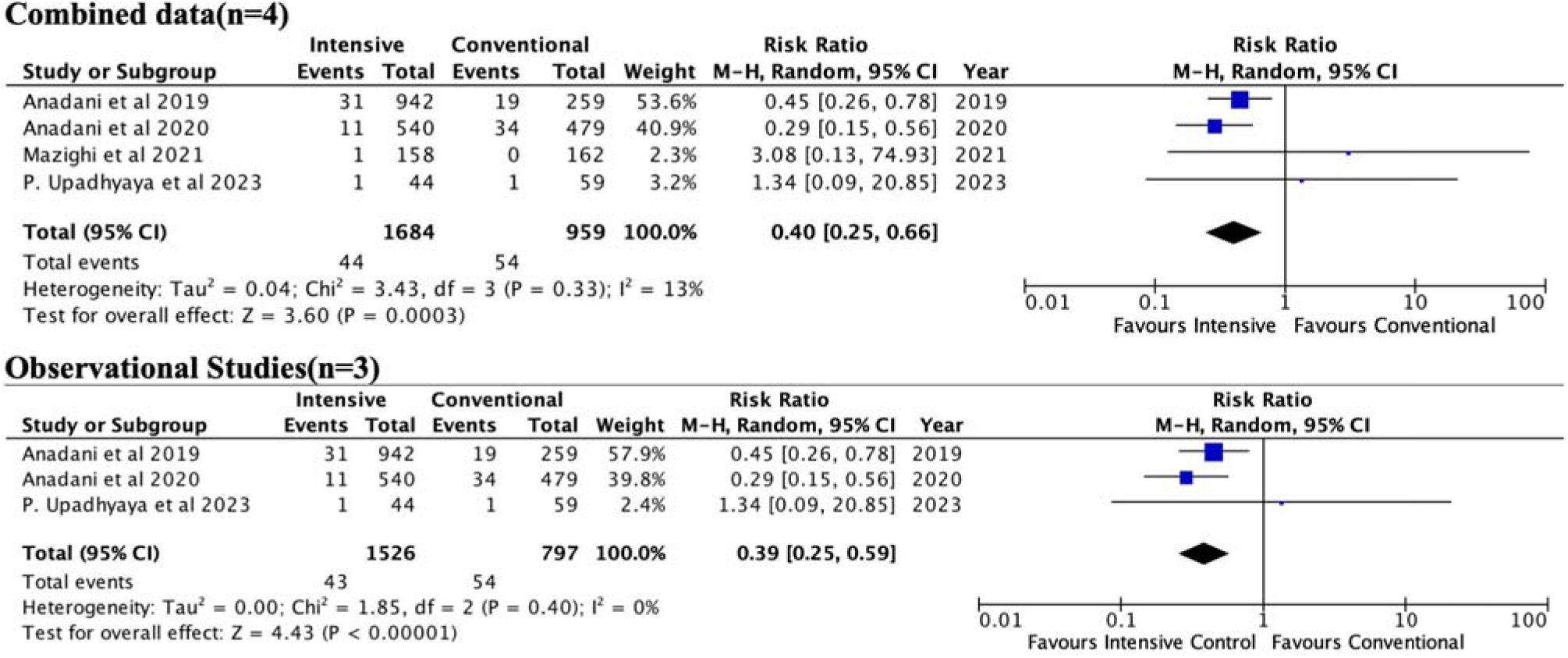
Forest plot for the outcome of decompressive surgery. The estimated risk ratio-RR is represented by a square and the corresponding 95% CI are shown for the outcome requirement of decompressive surgery. The square’s size is proportional to the study’s weight in the meta-analysis implying studies with larger squares have greater weight. The diamond beneath is the overall pooled estimate and its width denotes the 95%CI around the combined effect size.

A separate analysis of only observational study data from three studies showed that 43/1526 (2.82%) patients required decompressive surgery in the intensive arm compared to 54/797 (6.76%) patients in the conventional arm with the chances of needing a decompression higher in the conventional arm than intensive arm(RR-0.39; 95%CI-0.25-0.59; p<0.00001) as shown in Figure-5.^13,18,19^

### Optimal Systolic Blood Pressure cut-offs

We further explored the various SBP cut-offs (SBP at which anti-hypertensive medications would be started and the target SBP up to 24hours after EVT) used across the studies and segregated them into groups of studies using a cut-off of 140mmHg to define intensive control and 130mmHg to define intensive control.

### Primary outcome for a SBP cut-off of 140mmHg

Pooled analysis of seven studies(n=2156 patients in intensive arm and 1488 patients in conventional arm) for the primary outcome of mRS 0-2 at 90days with a cut-off of 140mmHg to define intensive control did not show in any significant differences between the intensive or less intensive/conventional control(RR-1.14; 95%CI-0.96-1.36; p=0.14) as shown in the Figure-6.^9–11,13,15–17^

**Figure-6.**
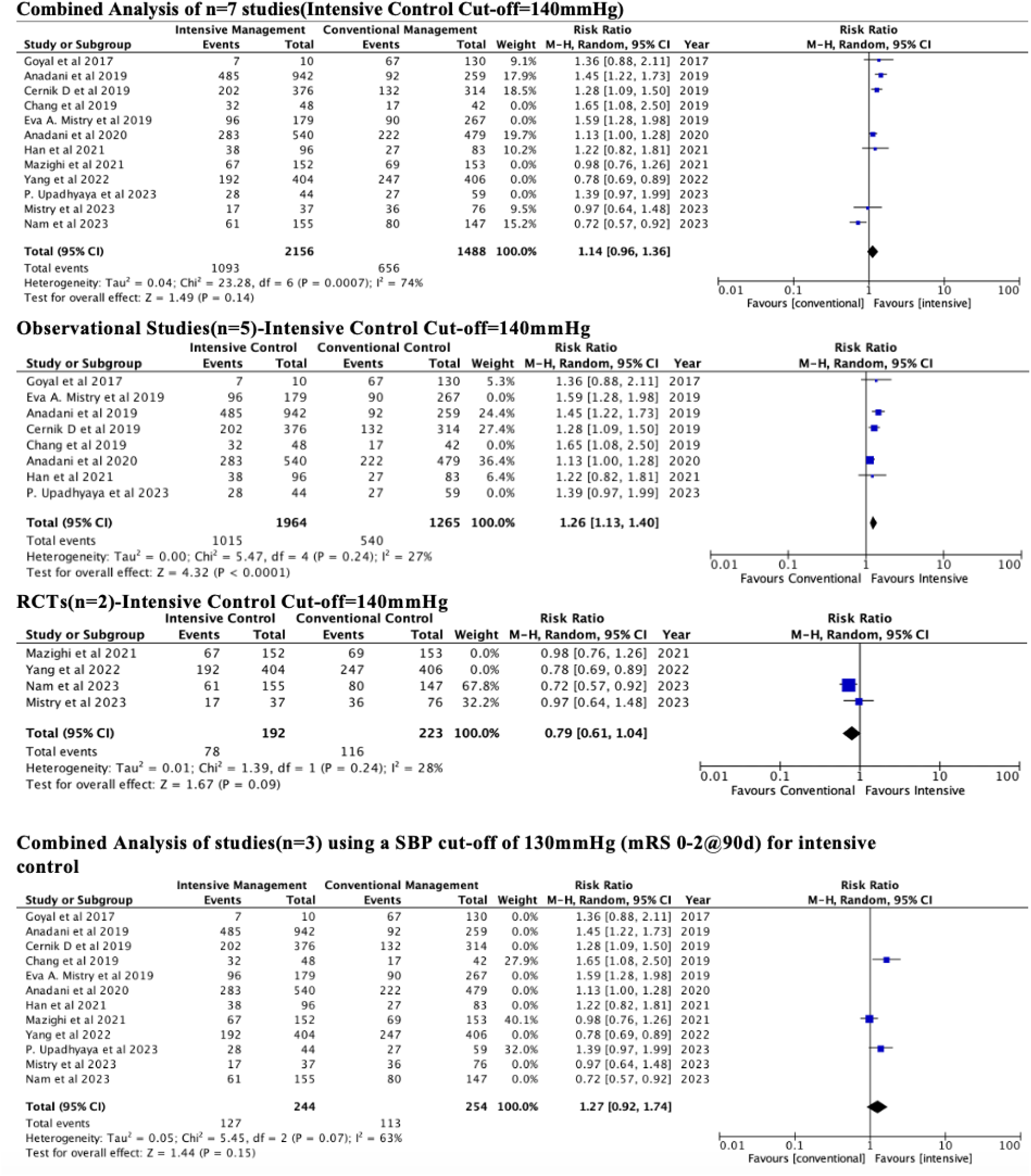
mRS 0-2 at 90 days with different SBP cut-off to define intensive control. The estimated risk ratio-RR is represented by a square and the corresponding 95% CI are shown for the primary outcome(mRS-0-2) at 90 days. The square’s size is proportional to the study’s weight in the meta-analysis implying studies with larger squares have greater weight. The diamond beneath is the overall pooled estimate and its width denotes the 95%CI around the combined effect size.

However a separate analysis of five observational studies (n=1964 patients in the intensive arm and 1265 patients in conventional arm) only using the 140mmHg cut-off for the primary outcome demonstrated a significant chance of favourable outcome in the intensive arm(RR-1.26; 95%CI-1.13-1.40; p<0.0001) as shown in Figure-6.^11,13,15–17^

A similar analysis for two RCTs (n=192 patients in intensive arm and 223 patients in the conventional arm) with the SBP cut-off of 140mmHg did not show a significant difference between the two groups(RR-0.79; 95%CI-0.61-1.04; p=0.09) as shown in Figure-6.^9,10^ Among the other outcomes death at 90days was significantly lower in the intensive control arm in the observational data involving five studies(n=1964 in intensive arm and n=1265 in conventional arm) with the cut-off of 140mmHg(RR-0.76; 95%CI-0.60-0.97; p=0.03) as shown in Supplemental Figure-2. The details about the other outcomes of sICH, decompressive surgery at different cut-offs can be found in the *Supplemental material-4 Figures-1,2,3*.

### Primary outcome for a SBP cut-off of 130mmHg

An analysis of three studies(n=244 in intensive arm and n=254 in conventional arm) with respect to the primary outcome using a target SBP cut-off of 130mmHg did not show any significant differences between the two groups(RR-1.27; 95%CI-0.92-1.74; p=0.15) as shown in Figure-6.^7,14,18^

The details about the other outcomes of sICH and decompressive surgery and different cut-offs can be found in the *Supplemental material-4;Figures-1,2,3*.

We could not explore other lower(one study) and higher cut-offs(two studies) due to limited number of studies in these groups and non-availability of individual patient data.

### Leave-out-one study Influence analysis

Leaving out the ENCHANTED-2/MT and OPTIMAL-BP results resulted in a change in the outcome of the pooled data such that the intensive arm had a higher odds of having a better functional outcome as compared to conventional BP management.^8,10^ Leave out one study analysis did not lead to any change in the primary outcome in a separate analysis of only observational studies. Similar to the combined analysis, leaving out the ENCHANTED-2/MT and OPTIMAL-BP results resulted in a change in the outcome of the RCT pooled data but the difference was such that neither arm was preferred^8,10^. Leaving out the other two RCTs did not change outcomes. The details are provided in the supplement *(Supplementary material-5)*.

### GRADE

The GRADEpro GDT (https://www.gradepro.org) was used to assess the certainty of evidence and was found to be moderate to low in case of observational studies and moderate in case of the RCTs. The summary of the GRADE findings is presented in the *Supplementary material-6*.

## Discussion

Our systematic review and meta-analysis involving eight observational studies and four RCTs analysed the effects of an intensive as compared to a conventional/less intensive blood pressure control up to 24 hours of thrombectomy in all strokes (eleven studies on anterior circulation and one on posterior circulation stroke). The results suggest that neither intensive nor conventional management of blood pressure post thrombectomy resulted in any difference in the functional outcomes i.e. mRS-0-2 at 90days. Similarly the risk of death and sICH were also similar in both groups. A significantly fewer patients required decompressive surgery in the intensive arm as compared to the conventional arm. Separate analysis of data from observational studies and data from RCTs revealed discordant results. The primary outcome i.e. achieving a mRS 0-2 at 90 days was higher in the intensive arm as compared to the conventional arm in observational studies whereas the conventional management was favoured from RCT data. Mortality at 90 days was also lower in the intensive arm as compared to the conventional arm in the observational data and both groups were comparable as per pooled data from the RCTs. Similarly the observational data favoured intensive control with lower rates of symptomatic ICH as compared to a conventional blood pressure control. The pooled data of the RCTs did not favour any particular arm. Decompressive surgery was more likely to be required in the conventional arm as compared to the intensive arm.

The optimal blood pressure after endovascular therapy has remained elusive ever since the procedure was introduced. Numerous hypotheses have been suggested favouring and negating intensive and conventional management. Arguments in favour of intensive management primarily derived from observational studies and stemmed from the fact that intensive control reduced the rates of symptomatic ICH, reduced cerebral hyper perfusion thereby reducing cerebral edema and improving outcomes.^13,19^ However with the robust RCTs like the ENCHANTED-2/BP-TARGET not favouring the intensive arm an alternate school of thought has emerged.^7,8^ Firstly, higher SBP might be associated with maintenance of perfusion to distal branches of the blood vessel after successful recanalization. A more intensive SBP control would lead to focal areas of hypoperfusion leading to infarct progression and poor outcomes. Secondly, a successful recanalization after EVT may have persistent venous postcapillary thrombosis and an intensive BP reduction could be detrimental and further promote infarct and ischemia progression.^20^

Blood pressure is a dynamic entity subject to cerebral autoregulation which in turn is unique to a patient. Blood pressure variability post EVT have also been observed to be associated with poor outcomes.^21^ It is therefore important to avoid wide fluctuations in the blood pressure.

The pooled data did not show any difference in outcomes with the type of BP management strategy used although there was a trend towards better outcomes in the intensive arm which could be due to the reduced risk of reperfusion injury and lower risk of haemorrhagic transformation.^11,16^ The requirement of decompressive surgery was also significantly lower in the intensive arm. This could also be explained by the fact that lower SBP probably reduced the risk of reperfusion injury and subsequent infarct expansion and cerebral edema.^16^ The separate analysis of observational and RCT data showed discordant results. This discordance in the observational and randomised data could be attributed to multiple factors. First, randomised trials are conducted in a controlled environment with strict BP targets and had patients for longer periods of time in the intended target. The higher rates of hypotensive events in the intensive group patients in the RCTs might have been a potential confounder for the poor outcomes in this group. On the other hand the BP in observational studies is more like the real world with some studies using the peak SBP and some using the mean SBP over a period of 24 hrs after the procedure. Since BP fluctuates throughout the day and the mean BP is unlikely to reflect these wide fluctuations and it is difficult to ascertain whether these so called ‘intensive’ arm patients were actually intensively treated.

Second, RCTs used drugs to both increase and decrease the blood pressure to set targets and it is possible that these drugs could have been a potential confounder with respect to unknown pleotropic effects of the anti-hypertensive drug.^8^ Such data was not available for all the observational studies.

Third, although the observational studies selected had a net low risk of bias (using the NOS tool) it is not possible to completely eliminate selection and observer’s bias inherent in all observational studies.

Fourth, the observational cut-offs used were different for different studies. We used the definition of intensive as defined by the study which is the reason for the significant heterogeneity. A total of five out of eight observational studies namely Goyal et al, Anadani et al(2019), Cernik et al, Anadani et al (2020) and Han et al used a SBP cut off of 140mmHg as a definition for intensive control.^13,15,17,19,22^ On the other hand 2 studies by Chang et al and P Upadhyay et al used a lower cut-off of 130mmHg.^14^ A SBP cut-off of 158mmHg was found to dichotomise functional outcomes in the Mistry et al (2019) study.^12^

The cut-offs used in the RCTs for defining intensive was also different across studies with the ENCHANTED-2/MT and BP-TARGET defining the upper limit of SBP for the intensive arm at 120mmHg and the BEST-II dividing the BP into three groups of 140,160 and 180 and the OPTIMAL-BP using a upper limit SBP of 140mmHg.^7–9^ Even though we did a separate analysis using different cut-offs of 140mmHg and 130mmHg to define the intensive arm there was no difference in the results as the observational data favoured the intensive arm. The difference was seen in the RCT data as removing ENCHANTED-2/MT and BP-TARGET trials resulted in no significant difference in the two arms.^7,8^ A lower cut off of 130mmHg also did not result in any significant differences between the arms. Access to individual patient data of these studies could have allowed us to study higher BP targets to identify the SBP sweet spot across studies by pooling the blood pressure recordings and subsequently dichotomising the outcomes.

Another important aspect to note is the heterogeneity in perfusion deficit of the patients recruited in the study. In the RCT data the ENCHANTED2/MT had reported a considerable number of patients with a significant perfusion deficit before the thrombectomy and such cases are extremely prone to reinfarction and infarct progression if the blood pressure is lowered.^8^

Further studies involving more patients and more uniform BP cut-offs will provide better answers in the future. The findings of our study based on RCT data which are considered Level-1 evidence favour a conventional management of blood pressure with target SBP of 140-180mmHg in line with the recommendations from the AHA/ASA.

## Strengths

The major strengths of this meta-analysis is the comprehensive search methodology and an inclusive study pooling data of all the published studies till date. It also includes data of one study with posterior circulation strokes.

## Limitations

The major limitation of this study is that it uses only data that was reported. A study with individual patient data could have led to better identification of the true cut-off SBP for achieving a good outcome and we could have explored higher targets in better detail. We also could not explore posterior circulation strokes separately as there was only one study exploring this.^17^ We could have performed meta-regression by adjusting for co-variates to predict whether age, recanalization grade or the admission blood pressure had an effect on the final outcome.

## Conclusion

Results of this meta-analysis suggest that combined data from all studies does not favour either intensive or conventional SBP control after EVT. Observational data showed intensive control to be associated with higher chances of functional recovery, reduced mortality and risk of symptomatic ICH. Data from RCTs showed conventional SBP control to be associated with improved functional outcomes but no effect on mortality suggesting conventional management as the preferred pragmatic approach. Further ongoing RCTs using homogenous SBP cut-offs will provide more clarity on the ideal SBP target after EVT.

## Data Availability

The data referred in the manuscript is copyright of the authors and should be appropriately.

## Acknowledgements

Nil

## Consent

Not required as this is a systematic review and meta-analysis.

## Ethical Statement

No ethical clearance was required as this is a systematic review and meta-analysis

## Contributions

Baikuntha Panigrahi: Drafting/revision of the manuscript for content, including medical writing for content; Major role in the acquisition of data; Study concept or design; Analysis or interpretation of data. Rohit Bhatia: Drafting/revision of the manuscript for content, including medical writing for content; Major role in the acquisition of data; Study concept or design; Analysis or interpretation of data. Partha Haldar: Drafting/revision of the manuscript for content, including medical writing for content; Analysis or interpretation of data. Risha Sarkar: Major role in the acquisition of data. Imnameren Longkumer: Major role in the acquisition of data.

## Supplemental Material(S)

**S1-**PRISMA checklist

**S2**-Review Question and PICO Statements

**S3**-Risk of Bias analysis

**S4**- Outcomes analysed using different SBP cut-offs

**S5**-Leave one study out analysis

**S6**-GRADE assessment of certainty of evidence

Supplemental Figures-S3

