## Supplemental File for "Optimal Systolic Blood pressure control after thrombectomy in acute ischemic stroke-a systematic review and meta-analysis"

### PRISMA 2020 Checklist

| Section and Topic | Item # | Checklist item | Location where item is reported |
|---|---|---|---|
| **TITLE** | | | |
| Title | 1 | Identify the report as a systematic review. | Yes<br>Page 1 Line 2 |
| **ABSTRACT** | | | |
| Abstract | 2 | See the PRISMA 2020 for Abstracts checklist. | Page 1 Line 4 |
| **INTRODUCTION** | | | |
| Rationale | 3 | Describe the rationale for the review in the context of existing knowledge. | Yes<br>Page 3 Line 3 |
| Objectives | 4 | Provide an explicit statement of the objective(s) or question(s) the review addresses. | Page 3 Line 26 and Supplement-2 with PICO statements |
| **METHODS** | | | |
| Eligibility criteria | 5 | Specify the inclusion and exclusion criteria for the review and how studies were grouped for the syntheses. | Page 4 Line 7 |
| Information sources | 6 | Specify all databases, registers, websites, organisations, reference lists and other sources searched or consulted to identify studies. Specify the date when each source was last searched or consulted. | Page 3 Line 44 |
| Search strategy | 7 | Present the full search strategies for all databases, registers and websites, including any filters and limits used. | Page 3 Line 45-50 |
| Selection process | 8 | Specify the methods used to decide whether a study met the inclusion criteria of the review, including how many reviewers screened each record and each report retrieved, whether they worked independently, and if applicable, details of automation tools used in the process. | Page 4 Line 5 |
| Data collection process | 9 | Specify the methods used to collect data from reports, including how many reviewers collected data from each report, whether they worked independently, any processes for obtaining or confirming data from study investigators, and if applicable, details of automation tools used in the process. | Page 4 Line 5 and Line 26 |
| Data items | 10a | List and define all outcomes for which data were sought. Specify whether all results that were compatible with each outcome domain in each study were sought (e.g. for all measures, time points, analyses), and if not, the methods used to decide which results to collect. | Page 4 Line 31-39 |
| | 10b | List and define all other variables for which data were sought (e.g. participant and intervention characteristics, funding sources). Describe any assumptions made about any missing or unclear information. | Page 4 Line 31 |
| Study risk of bias assessment | 11 | Specify the methods used to assess risk of bias in the included studies, including details of the tool(s) used, how many reviewers assessed each study and whether they worked independently, and if applicable, details of automation tools used in the process. | Page 4 Line 42 |
| Effect measures | 12 | Specify for each outcome the effect measure(s) (e.g. risk ratio, mean difference) used in the synthesis or presentation of results. | Page 5 Line 1-2 |
| Synthesis methods | 13a | Describe the processes used to decide which studies were eligible for each synthesis (e.g. tabulating the study intervention characteristics and comparing against the planned groups for each synthesis (item #5)). | Page 4 Line 15 |
| | 13b | Describe any methods required to prepare the data for presentation or synthesis, such as handling of missing summary statistics, or data conversions. | Page 4 Line 31-39 |
| | 13c | Describe any methods used to tabulate or visually display results of individual studies and syntheses. | Page 5 Line 35 |

PRISMA 2020 Checklist

| Section and Topic | Item # | Checklist item | Location where item is reported |
|---|---|---|---|
| | 13d | Describe any methods used to synthesize results and provide a rationale for the choice(s). If meta-analysis was performed, describe the model(s), method(s) to identify the presence and extent of statistical heterogeneity, and software package(s) used. | Page 4 Line 47 |
| | 13e | Describe any methods used to explore possible causes of heterogeneity among study results (e.g. subgroup analysis, meta-regression). | Page 5 Line-1 |
| | 13f | Describe any sensitivity analyses conducted to assess robustness of the synthesized results. | Page 5 line 2 |
| Reporting bias assessment | 14 | Describe any methods used to assess risk of bias due to missing results in a synthesis (arising from reporting biases). | Page 4 Line 41 |
| Certainty assessment | 15 | Describe any methods used to assess certainty (or confidence) in the body of evidence for an outcome. | GRADE-Page 5 Line-3 |
| **RESULTS** | | | |
| Study selection | 16a | Describe the results of the search and selection process, from the number of records identified in the search to the number of studies included in the review, ideally using a flow diagram. | Page 5 Line 7 |
| | 16b | Cite studies that might appear to meet the inclusion criteria, but which were excluded, and explain why they were excluded. | Page 5 line 12 |
| Study characteristics | 17 | Cite each included study and present its characteristics. | Page 5 Line 15 |
| Risk of bias in studies | 18 | Present assessments of risk of bias for each included study. | Page 5 Line 32 |
| Results of individual studies | 19 | For all outcomes, present, for each study: (a) summary statistics for each group (where appropriate) and (b) an effect estimate and its precision (e.g. confidence/credible interval), ideally using structured tables or plots. | Page-5 Line 44 Figures-2,3,4,5,6 |
| Results of syntheses | 20a | For each synthesis, briefly summarise the characteristics and risk of bias among contributing studies. | Supplementary material-3 |
| | 20b | Present results of all statistical syntheses conducted. If meta-analysis was done, present for each the summary estimate and its precision (e.g. confidence/credible interval) and measures of statistical heterogeneity. If comparing groups, describe the direction of the effect. | Figures 2-6 |
| | 20c | Present results of all investigations of possible causes of heterogeneity among study results. | Page 5 Line 15 |
| | 20d | Present results of all sensitivity analyses conducted to assess the robustness of the synthesized results. | Page 7 Line 40 |
| Reporting biases | 21 | Present assessments of risk of bias due to missing results (arising from reporting biases) for each synthesis assessed. | Supplementary material-3 |
| Certainty of evidence | 22 | Present assessments of certainty (or confidence) in the body of evidence for each outcome assessed. | Supplement-6 |
| **DISCUSSION** | | | |
| Discussion | 23a | Provide a general interpretation of the results in the context of other evidence. | Page 8 line 27-39 |
| | 23b | Discuss any limitations of the evidence included in the review. | Page 9 Line 1 |
| | 23c | Discuss any limitations of the review processes used. | Page 10 line 9 |
| | 23d | Discuss implications of the results for practice, policy, and future research. | Page 10 Line |

### PRISMA 2020 Checklist

| Section and Topic | Item # | Checklist item | Location where item is reported |
|---|---|---|---|
| | | | 20 |
| **OTHER INFORMATION** | | | |
| Registration and protocol | 24a | Provide registration information for the review, including register name and registration number, or state that the review was not registered. | Page 3 Line 41 |
| | 24b | Indicate where the review protocol can be accessed, or state that a protocol was not prepared. | Page 3 Line 40 |
| | 24c | Describe and explain any amendments to information provided at registration or in the protocol. | Page 3 Line 41 |
| Support | 25 | Describe sources of financial or non-financial support for the review, and the role of the funders or sponsors in the review. | Page 11 Line 4 |
| Competing interests | 26 | Declare any competing interests of review authors. | Page 11 Line 6 |
| Availability of data, code and other materials | 27 | Report which of the following are publicly available and where they can be found: template data collection forms; data extracted from included studies; data used for all analyses; analytic code; any other materials used in the review. | Supplementary material 1-6 |

*From:* Page MJ, McKenzie JE, Bossuyt PM, Boutron I, Hoffmann TC, Mulrow CD, et al. The PRISMA 2020 statement: an updated guideline for reporting systematic reviews. BMJ 2021;372:n71. doi: 10.1136/bmj.n71

For more information, visit: http://www.prisma-statement.org/

### Supplementary material 2

**Review Question**

In adult patients with an acute ischemic stroke due to a large vessel occlusion who undergo thrombectomy does an intensive blood pressure control (Target SBP cutoff as defined by the respective study or upper limit of target SBP cut-off of 140mmHg) as compared to a less intensive blood pressure control up to 24 hours after the procedure lead to a good functional outcome?

**Table1-PICO Statements**

| | |
|---|---|
| **Population** | All adult (>18years) patients of acute ischemic stroke due to a large vessel occlusion undergoing thrombectomy. |
| **Intervention** | Intensive Blood Pressure Control (Target SBP cutoff as defined in the respective study definition) up to 24 hours after the procedure. |
| **Control** | Less Intensive Blood Pressure Control (Target SBP cutoff as defined by the respective study definition) up to 24 hours after the procedure. |
| **Primary Efficacy Outcome** | Proportion of patients achieving functional independence at 90 days defined as mRS of 0-2. |

SBP-Systolic Blood Pressure; mRS-modified Rankin Scale

**Safety outcomes** assessed-

1.Death at 90days

2.Requirement of decompressive surgery

3.Symptomatic ICH as per respective study definition.

### Supplement-3

Risk of Bias

Table-1-New Castle Ottawa Scale

| | | | Selection | | | | Comp | Outcome | | | | | |
|---|---|---|---|---|---|---|---|---|---|---|---|---|---|
| Sl No | Author | Year | Repr | Selec | Ascert | Demor | Comp | Assess | Was fol | Adequ | Total Stars | Total Stars cc | Impression |
| 1 | Goyal et al | 2017 | * | * | * | * | ** | * | * | * | ********* | 9 | Good |
| 2 | Eva A. Mistr | 2019 | * | * | * | * | * | * | * | * | ******** | 8 | Good |
| 3 | Anadani et al | 2019 | * | * | * | * | * | 0 | * | * | ******** | 7 | Good |
| 4 | Chang et al | 2019 | * | * | * | * | ** | * | * | * | ********* | 9 | Good |
| 5 | Cernik D et a | 2019 | * | * | * | * | ** | 0 | * | * | ******** | 8 | Good |
| 6 | Anadani et al | 2020 | * | * | * | * | ** | 0 | * | * | ******** | 8 | Good |
| 7 | Han et al | 2021 | * | * | * | * | ** | * | * | * | ********* | 9 | Good |
| 8 | P. Upadhyay | 2023 | * | * | * | * | ** | 0 | * | * | ******** | 8 | Good |
| | | | | | | | | | | | | 8.25 | |
| | | | | | | | | | | | | 8 | |

### Table-2-ROB2 tool for the studies included

| | | | | | |
|---|---|---|---|---|---|
| Basic informa | Time | 2024/03/02 20.36 | | | |
| | Unique ID | 1 | 2 | 3 | 4 |
| | Assessor | Dr.Baikuntha | Dr.Baikuntha | Dr.Baikuntha | Dr.Baikuntha |
| | Study ID | Yang 2022 | Mazighi 2021 | Nam 2023 | Mistry 2023 |
| | Reference | Yang 2022 | Mazighi 2021 | Nam 2023 | Mistry 2023 |
| | Experimental | Intensive Control | Intensive Control | Intensive Control | Intensive Control |
| | Comparator | Conventional | Conventional | Conventional | Conventional |
| | Outcome | mRS 0-2 | mRS 0-2 | mRS 0-2 | mRS 0-2 |
| | Results | | | | |
| | Aim | assignment to intervention (the 'intentio | assignment to intervention (the 'intention | assignment to intervention (the 'inten | assignment to intervention (the 'intention-to |
| | Effect of adhering to intervention? | NA | NA | NA | NA |
| | Weight | | | | |
| Domain 1. R | Sources | Journal article(s); Trial protocol; Statist | Journal article(s); Trial protocol; Statistic | Journal article(s); Trial protocol; Sta | Journal article(s); Trial protocol; Statistica |
| | 1.1 | Y | Y | Y | Y |
| | 1.2 | Y | Y | Y | Y |
| | Note for 1.1&1.2 | | | | |
| | 1.3 | N | N | N | N |
| | Note for 1.3 | | | | |
| | 1.0 Algorithm result | | | | |
| | 1.0 Assessor's Judgement | Low | Low | Low | Low |
| Domain 2. D | 1.0 General note | | | | |
| | 1.0 Optional Question | | | | |
| | 1.0 Note for optional question | | | | |
| | 2.1 | Y | Y | Y | Y |
| | 2.2 | Y | Y | Y | Y |
| | Note for 2.1&2.2 | | | | |
| | 2.3 | PY | Y | Y | Y |
| | Note for 2.3 | | | | |
| | 2.4 | PY | Y | Y | Y |
| | Note for 2.4 | | | | |
| | 2.5 | PY | PY | PY | PY |
| | Note for 2.5 | | | | |
| | 2.6 | Y | N | N | N |
| | Note for 2.6 | | | | |
| Domain 3. M | 2.7 | NA | | | |
| | Note for 2.7 | | | | |
| | 2.0 Algorithm result | | | | |
| | 2.0 Assessor's Judgement | Some concerns | High | High | High |
| | 2.0 General Notes | | | | |
| | 2.0 Optional Question | | | | |
| | 2.0 Note for optional question | | | | |
| | 3.1 | Y | Y | Y | Y |
| | Note for 3.1 | | | | |
| Domain 4. M | 3.2 | NA | | | |
| | Note for 3.2 | | | | |
| | 3.3 | NA | | | |
| | Note for 3.3&3.4 | | | | |
| | 3.4 | NA | | | |
| | 3.0 Algorithm result | | | | |
| | 3.0 Assessor's judgement | Low | Low | Low | Low |
| | 3.0 Gerenal notes | | | | |
| | 3.0 Optional Question | | | | |
| | 3.0 Note for optional question | | | | |
| | 4.1 | N | N | N | N |
| Domain 5. S | Note for 4.1 | | | | |
| | 4.2 | N | N | N | N |
| | Note for 4.2 | | | | |
| | 4.3 | N | N | N | N |
| | Note for 4.3 | | | | |
| | 4.4 | NA | | | |
| | Note for 4.4&4.5 | | | | |
| | 4.5 | NA | | | |
| Domain 6. O | 4.0 Algorithm result | | | | |
| | 4.0 Assessor's Judgement | Low | Low | Low | Low |
| | 4.0 General note | | | | |
| | 4.0 Optional Question | | | | |
| | 4.0 Note for optional question | | | | |
| | 5.1 | Y | Y | Y | Y |
| | Note for 5.1 | | | | |
| | 5.2 | N | N | N | N |
| | Note for 5.2 | | | | |
| | 5.3 | N | N | N | N |
| | Note for 5.3 | | | | |
| | 5.0 Algorithm result | | | | |
| | 5.0 Assessor's Judgement | Low | Low | Low | Low |
| | 5.0 General note | | | | |
| | 5.0 Optional Question | | | | |
| | 5.0 Note for optional question | | | | |
| | Algorithm's overall Judgement | | | | |
| | Assessor's overall Judgement | Some concerns | High risk | High risk | High risk |

#### Supplement-4

**Supplemental Figure-1-Death at 90 days using data from studies using different SBP targets to define intensive control**

##### Combined analysis(n=7) of studies with SBP cut-off of 140mmHg to define intensive control

| Study or Subgroup | Intensive Events | Intensive Total | Conventional Events | Conventional Total | Weight | Risk Ratio M-H, Random, 95% CI | Year |
|---|---|---|---|---|---|---|---|
| Goyal et al 2017 | 0 | 10 | 30 | 130 | 0.6% | 0.20 [0.01, 2.98] | 2017 |
| Cernik D et al 2019 | 88 | 376 | 101 | 314 | 27.0% | 0.73 [0.57, 0.93] | 2019 |
| Chang et al 2019 | 5 | 48 | 6 | 42 | 0.0% | 0.73 [0.24, 2.22] | 2019 |
| Eva A. Mistry et al 2019 | 20 | 179 | 59 | 267 | 0.0% | 0.51 [0.32, 0.81] | 2019 |
| Anadani et al 2019 | 150 | 942 | 67 | 259 | 26.2% | 0.62 [0.48, 0.79] | 2019 |
| Anadani et al 2020 | 84 | 540 | 91 | 479 | 25.0% | 0.82 [0.63, 1.07] | 2020 |
| Han et al 2021 | 21 | 96 | 12 | 83 | 8.9% | 1.51 [0.79, 2.89] | 2021 |
| Mazighi et al 2021 | 29 | 152 | 21 | 153 | 0.0% | 1.39 [0.83, 2.33] | 2021 |
| Yang et al 2022 | 66 | 406 | 61 | 408 | 0.0% | 1.09 [0.79, 1.50] | 2022 |
| Mistry et al 2023 | 7 | 37 | 20 | 76 | 6.8% | 0.72 [0.33, 1.55] | 2023 |
| Nam et al 2023 | 12 | 155 | 8 | 147 | 5.5% | 1.42 [0.60, 3.38] | 2023 |
| P. Upadhyaya et al 2023 | 5 | 44 | 9 | 59 | 0.0% | 0.74 [0.27, 2.07] | 2023 |
| **Total (95% CI)** | | **2156** | | **1488** | **100.0%** | **0.79 [0.63, 0.98]** | |
| Total events | 362 | | 329 | | | | |

Heterogeneity: $Tau^2 = 0.03$; $Chi^2 = 10.42$, df = 6 (P = 0.11); $I^2 = 42\%$
Test for overall effect: Z = 2.15 (P = 0.03)

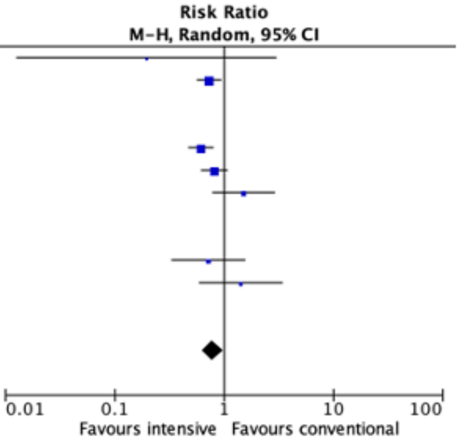


##### Observational studies(n=5) with SBP cut-off of 140mmHg to define intensive control

| Study or Subgroup | Intensive Arm Events | Intensive Arm Total | Conventional Arm Events | Conventional Arm Total | Weight | Risk Ratio M-H, Random, 95% CI | Year |
|---|---|---|---|---|---|---|---|
| Goyal et al 2017 | 0 | 10 | 30 | 130 | 0.7% | 0.20 [0.01, 2.98] | 2017 |
| Eva A. Mistry et al 2019 | 20 | 179 | 59 | 267 | 0.0% | 0.51 [0.32, 0.81] | 2019 |
| Anadani et al 2019 | 150 | 942 | 67 | 259 | 29.8% | 0.62 [0.48, 0.79] | 2019 |
| Cernik D et al 2019 | 88 | 376 | 101 | 314 | 30.6% | 0.73 [0.57, 0.93] | 2019 |
| Chang et al 2019 | 5 | 48 | 6 | 42 | 0.0% | 0.73 [0.24, 2.22] | 2019 |
| Anadani et al 2020 | 84 | 540 | 91 | 479 | 28.5% | 0.82 [0.63, 1.07] | 2020 |
| Han et al 2021 | 21 | 96 | 12 | 83 | 10.4% | 1.51 [0.79, 2.89] | 2021 |
| P. Upadhyaya et al 2023 | 5 | 44 | 9 | 59 | 0.0% | 0.74 [0.27, 2.07] | 2023 |
| **Total (95% CI)** | | **1964** | | **1265** | **100.0%** | **0.76 [0.60, 0.97]** | |
| Total events | 343 | | 301 | | | | |

Heterogeneity: $Tau^2 = 0.03$; $Chi^2 = 8.23$, df = 4 (P = 0.08); $I^2 = 51\%$
Test for overall effect: Z = 2.21 (P = 0.03)

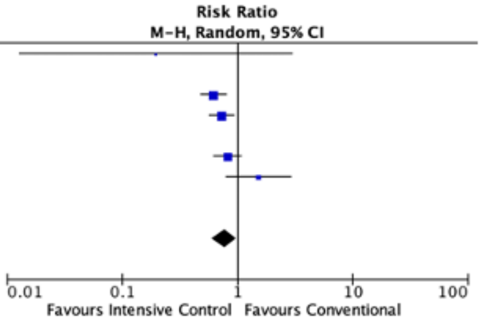


##### RCTs(n=2) with SBP cut-off of 140mmHg to define intensive control

| Study or Subgroup | Intensive Control Events | Intensive Control Total | Conventional Control Events | Conventional Control Total | Weight | Risk Ratio M-H, Random, 95% CI | Year |
|---|---|---|---|---|---|---|---|
| Mazighi et al 2021 | 29 | 152 | 21 | 153 | 0.0% | 1.39 [0.83, 2.33] | 2021 |
| Yang et al 2022 | 66 | 406 | 61 | 408 | 0.0% | 1.09 [0.79, 1.50] | 2022 |
| Nam et al 2023 | 12 | 155 | 8 | 147 | 45.5% | 1.42 [0.60, 3.38] | 2023 |
| Mistry et al 2023 | 7 | 37 | 20 | 76 | 54.5% | 0.72 [0.33, 1.55] | 2023 |
| **Total (95% CI)** | | **192** | | **223** | **100.0%** | **0.98 [0.50, 1.91]** | |
| Total events | 19 | | 28 | | | | |

Heterogeneity: $Tau^2 = 0.06$; $Chi^2 = 1.34$, df = 1 (P = 0.25); $I^2 = 26\%$
Test for overall effect: Z = 0.06 (P = 0.95)

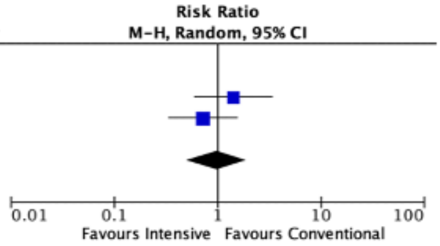


##### Combined Analysis of studies(n=3) using a SBP cut-off of 130mmHg (Death@90d) for intensive control

| Study or Subgroup | Intensive Events | Intensive Total | Conventional Events | Conventional Total | Weight | Risk Ratio M-H, Random, 95% CI | Year |
|---|---|---|---|---|---|---|---|
| Goyal et al 2017 | 0 | 10 | 30 | 130 | 0.0% | 0.20 [0.01, 2.98] | 2017 |
| Cernik D et al 2019 | 88 | 376 | 101 | 314 | 0.0% | 0.73 [0.57, 0.93] | 2019 |
| Chang et al 2019 | 5 | 48 | 6 | 42 | 14.6% | 0.73 [0.24, 2.22] | 2019 |
| Eva A. Mistry et al 2019 | 20 | 179 | 59 | 267 | 0.0% | 0.51 [0.32, 0.81] | 2019 |
| Anadani et al 2019 | 150 | 942 | 67 | 259 | 0.0% | 0.62 [0.48, 0.79] | 2019 |
| Anadani et al 2020 | 84 | 540 | 91 | 479 | 0.0% | 0.82 [0.63, 1.07] | 2020 |
| Han et al 2021 | 21 | 96 | 12 | 83 | 0.0% | 1.51 [0.79, 2.89] | 2021 |
| Mazighi et al 2021 | 29 | 152 | 21 | 153 | 68.1% | 1.39 [0.83, 2.33] | 2021 |
| Yang et al 2022 | 66 | 406 | 61 | 408 | 0.0% | 1.09 [0.79, 1.50] | 2022 |
| Mistry et al 2023 | 7 | 37 | 20 | 76 | 0.0% | 0.72 [0.33, 1.55] | 2023 |
| Nam et al 2023 | 12 | 155 | 8 | 147 | 0.0% | 1.42 [0.60, 3.38] | 2023 |
| P. Upadhyaya et al 2023 | 5 | 44 | 9 | 59 | 17.3% | 0.74 [0.27, 2.07] | 2023 |
| **Total (95% CI)** | | **244** | | **254** | **100.0%** | **1.14 [0.74, 1.74]** | |
| Total events | 39 | | 36 | | | | |

Heterogeneity: $Tau^2 = 0.00$; $Chi^2 = 1.86$, df = 2 (P = 0.39); $I^2 = 0\%$
Test for overall effect: Z = 0.59 (P = 0.56)

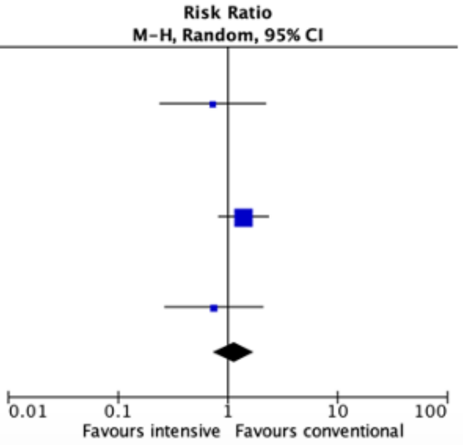

**Supplemental Figure-2**
**Symptomatic intracranial haemorrhage(sICH) using data from studies using different SBP targets to define intensive control**

##### Combined Analysis(n=6) for studies using a SBP cut-off of 140mmHg for intensive control

| Study or Subgroup | Intensive Events | Intensive Total | Conventional Events | Conventional Total | Weight | Risk Ratio M-H, Random, 95% CI | Year |
|---|---|---|---|---|---|---|---|
| Goyal et al 2017 | 1 | 10 | 7 | 130 | 3.4% | 1.86 [0.25, 13.64] | 2017 |
| Anadani et al 2019 | 37 | 942 | 20 | 259 | 27.0% | 0.51 [0.30, 0.86] | 2019 |
| Anadani et al 2020 | 16 | 540 | 29 | 479 | 23.5% | 0.49 [0.27, 0.89] | 2020 |
| Han et al 2021 | 19 | 96 | 16 | 83 | 23.6% | 1.03 [0.57, 1.86] | 2021 |
| Mazighi et al 2021 | 17 | 154 | 11 | 157 | 0.0% | 1.58 [0.76, 3.25] | 2021 |
| Yang et al 2022 | 23 | 407 | 25 | 409 | 0.0% | 0.92 [0.53, 1.60] | 2022 |
| Nam et al 2023 | 14 | 155 | 12 | 149 | 18.1% | 1.12 [0.54, 2.34] | 2023 |
| P. Upadhyaya et al 2023 | 1 | 44 | 4 | 59 | 0.0% | 0.34 [0.04, 2.90] | 2023 |
| Mistry et al 2023 | 2 | 37 | 3 | 72 | 4.4% | 1.30 [0.23, 7.43] | 2023 |
| **Total (95% CI)** | | **1780** | | **1172** | **100.0%** | **0.75 [0.51, 1.10]** | |
| Total events | 89 | | 87 | | | | |

Heterogeneity: Tau² = 0.07; Chi² = 7.33, df = 5 (P = 0.20); I² = 32%
Test for overall effect: Z = 1.49 (P = 0.14)

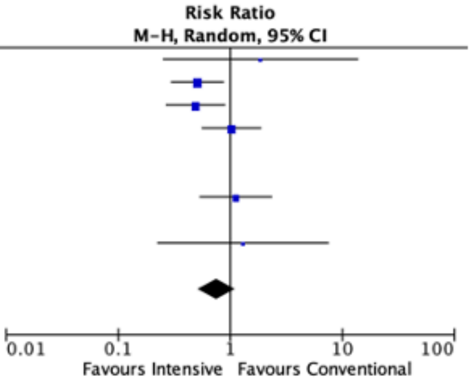


##### Observational studies(n=4)

| Study or Subgroup | Intensive Control Events | Intensive Control Total | Conventional Events | Conventional Total | Weight | Risk Ratio M-H, Random, 95% CI | Year |
|---|---|---|---|---|---|---|---|
| Goyal et al 2017 | 1 | 10 | 7 | 130 | 4.8% | 1.86 [0.25, 13.64] | 2017 |
| Anadani et al 2019 | 37 | 942 | 20 | 259 | 34.5% | 0.51 [0.30, 0.86] | 2019 |
| Anadani et al 2020 | 16 | 540 | 29 | 479 | 30.4% | 0.49 [0.27, 0.89] | 2020 |
| Han et al 2021 | 19 | 96 | 16 | 83 | 30.4% | 1.03 [0.57, 1.86] | 2021 |
| P. Upadhyaya et al 2023 | 1 | 44 | 4 | 59 | 0.0% | 0.34 [0.04, 2.90] | 2023 |
| **Total (95% CI)** | | **1588** | | **951** | **100.0%** | **0.66 [0.42, 1.04]** | |
| Total events | 73 | | 72 | | | | |

Heterogeneity: Tau² = 0.08; Chi² = 5.03, df = 3 (P = 0.17); I² = 40%
Test for overall effect: Z = 1.79 (P = 0.07)

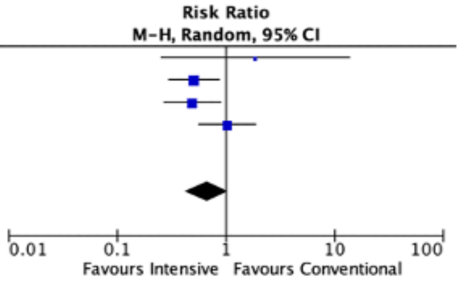


##### RCTs(n=2)

| Study or Subgroup | Intensive Arm Events | Intensive Arm Total | Conventional Arm Events | Conventional Arm Total | Weight | Risk Ratio M-H, Random, 95% CI | Year |
|---|---|---|---|---|---|---|---|
| Mazighi et al 2021 | 17 | 154 | 11 | 157 | 0.0% | 1.58 [0.76, 3.25] | 2021 |
| Yang et al 2022 | 23 | 407 | 25 | 409 | 0.0% | 0.92 [0.53, 1.60] | 2022 |
| Nam et al 2023 | 14 | 155 | 12 | 149 | 84.8% | 1.12 [0.54, 2.34] | 2023 |
| Mistry et al 2023 | 2 | 37 | 3 | 72 | 15.2% | 1.30 [0.23, 7.43] | 2023 |
| **Total (95% CI)** | | **192** | | **221** | **100.0%** | **1.15 [0.58, 2.26]** | |
| Total events | 16 | | 15 | | | | |

Heterogeneity: Tau² = 0.00; Chi² = 0.02, df = 1 (P = 0.88); I² = 0%
Test for overall effect: Z = 0.39 (P = 0.69)

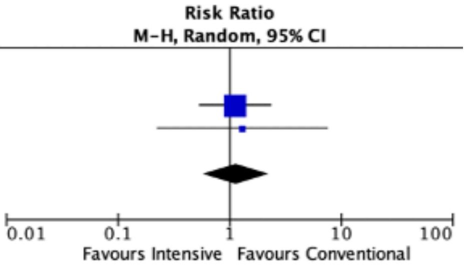


##### Combined Analysis of studies(n=2) using a SBP cut-off of 130mmHg (sICH) for intensive control

| Study or Subgroup | Intensive Events | Intensive Total | Conventional Events | Conventional Total | Weight | Risk Ratio M-H, Random, 95% CI | Year |
|---|---|---|---|---|---|---|---|
| Goyal et al 2017 | 1 | 10 | 7 | 130 | 0.0% | 1.86 [0.25, 13.64] | 2017 |
| Anadani et al 2019 | 37 | 942 | 20 | 259 | 0.0% | 0.51 [0.30, 0.86] | 2019 |
| Anadani et al 2020 | 16 | 540 | 29 | 479 | 0.0% | 0.49 [0.27, 0.89] | 2020 |
| Han et al 2021 | 19 | 96 | 16 | 83 | 0.0% | 1.03 [0.57, 1.86] | 2021 |
| Mazighi et al 2021 | 17 | 154 | 11 | 157 | 72.1% | 1.58 [0.76, 3.25] | 2021 |
| Yang et al 2022 | 23 | 407 | 25 | 409 | 0.0% | 0.92 [0.53, 1.60] | 2022 |
| Nam et al 2023 | 14 | 155 | 12 | 149 | 0.0% | 1.12 [0.54, 2.34] | 2023 |
| P. Upadhyaya et al 2023 | 1 | 44 | 4 | 59 | 27.9% | 0.34 [0.04, 2.90] | 2023 |
| Mistry et al 2023 | 2 | 37 | 3 | 72 | 0.0% | 1.30 [0.23, 7.43] | 2023 |
| **Total (95% CI)** | | **198** | | **216** | **100.0%** | **1.02 [0.26, 4.02]** | |
| Total events | 18 | | 15 | | | | |

Heterogeneity: Tau² = 0.54; Chi² = 1.80, df = 1 (P = 0.18); I² = 44%
Test for overall effect: Z = 0.03 (P = 0.97)

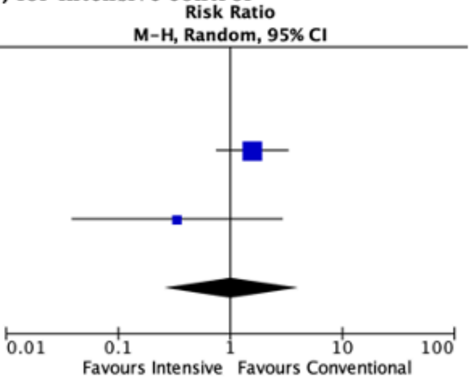

**Supplemental Figure-3**
**Decompressive surgery using data from studies using different SBP targets to define intensive control**

**Combined Analysis(n=2) for studies using a SBP cut-off of 140mmHg for intensive control**

| Study or Subgroup | Intensive Events | Intensive Total | Conventional Events | Conventional Total | Weight | Risk Ratio M-H, Random, 95% CI | Year |
|---|---|---|---|---|---|---|---|
| Anadani et al 2019 | 31 | 942 | 19 | 259 | 58.9% | 0.45 [0.26, 0.78] | 2019 |
| Anadani et al 2020 | 11 | 540 | 34 | 479 | 41.1% | 0.29 [0.15, 0.56] | 2020 |
| Mazighi et al 2021 | 1 | 158 | 0 | 162 | 0.0% | 3.08 [0.13, 74.93] | 2021 |
| P. Upadhyaya et al 2023 | 1 | 44 | 1 | 59 | 0.0% | 1.34 [0.09, 20.85] | 2023 |
| **Total (95% CI)** | | **1482** | | **738** | **100.0%** | **0.37 [0.24, 0.58]** | |
| Total events | 42 | | 53 | | | | |

Heterogeneity: $Tau^2 = 0.00$; $Chi^2 = 1.05$, df = 1 (P = 0.31); $I^2 = 4\%$
Test for overall effect: Z = 4.42 (P < 0.00001)

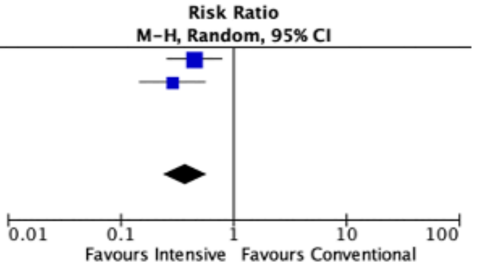


**Combined Analysis(n=2) for studies using a SBP cut-off of 130mmHg for intensive control**

| Study or Subgroup | Intensive Events | Intensive Total | Conventional Events | Conventional Total | Weight | Risk Ratio M-H, Random, 95% CI | Year |
|---|---|---|---|---|---|---|---|
| Anadani et al 2019 | 31 | 942 | 19 | 259 | 0.0% | 0.45 [0.26, 0.78] | 2019 |
| Anadani et al 2020 | 11 | 540 | 34 | 479 | 0.0% | 0.29 [0.15, 0.56] | 2020 |
| Mazighi et al 2021 | 1 | 158 | 0 | 162 | 42.5% | 3.08 [0.13, 74.93] | 2021 |
| P. Upadhyaya et al 2023 | 1 | 44 | 1 | 59 | 57.5% | 1.34 [0.09, 20.85] | 2023 |
| **Total (95% CI)** | | **202** | | **221** | **100.0%** | **1.91 [0.24, 15.29]** | |
| Total events | 2 | | 1 | | | | |

Heterogeneity: $Tau^2 = 0.00$; $Chi^2 = 0.15$, df = 1 (P = 0.70); $I^2 = 0\%$
Test for overall effect: Z = 0.61 (P = 0.54)

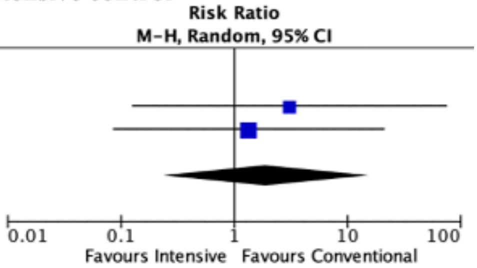

### Supplement-2-Leave out one study analysis

Studies in **bold** and having a * had a significant change in the primary outcome after leaving them out.

#### Leave out Goyal et al 2017

| | Intensive Management | | Conventional Management | | | Risk Ratio | |
|---|---|---|---|---|---|---|---|
| Study or Subgroup | Events | Total | Events | Total | Weight | M-H, Random, 95% CI | Year |
| Goyal et al 2017 | 7 | 10 | 67 | 130 | 0.0% | 1.36 [0.88, 2.11] | 2017 |
| Anadani et al 2019 | 485 | 942 | 92 | 259 | 10.4% | 1.45 [1.22, 1.73] | 2019 |
| Cernik D et al 2019 | 202 | 376 | 132 | 314 | 10.6% | 1.28 [1.09, 1.50] | 2019 |
| Chang et al 2019 | 32 | 48 | 17 | 42 | 6.9% | 1.65 [1.08, 2.50] | 2019 |
| Eva A. Mistry et al 2019 | 96 | 179 | 90 | 267 | 9.8% | 1.59 [1.28, 1.98] | 2019 |
| Anadani et al 2020 | 283 | 540 | 222 | 479 | 10.9% | 1.13 [1.00, 1.28] | 2020 |
| Han et al 2021 | 38 | 96 | 27 | 83 | 7.2% | 1.22 [0.82, 1.81] | 2021 |
| Mazighi et al 2021 | 67 | 152 | 69 | 153 | 9.3% | 0.98 [0.76, 1.26] | 2021 |
| Yang et al 2022 | 192 | 404 | 247 | 406 | 10.9% | 0.78 [0.69, 0.89] | 2022 |
| P. Upadhyaya et al 2023 | 28 | 44 | 27 | 59 | 7.8% | 1.39 [0.97, 1.99] | 2023 |
| Mistry et al 2023 | 17 | 37 | 36 | 76 | 6.8% | 0.97 [0.64, 1.48] | 2023 |
| Nam et al 2023 | 61 | 155 | 80 | 147 | 9.4% | 0.72 [0.57, 0.92] | 2023 |
| **Total (95% CI)** | | **2973** | | **2285** | **100.0%** | **1.15 [0.96, 1.36]** | |
| Total events | 1501 | | 1039 | | | | |

Heterogeneity: Tau$^2$ = 0.07; Chi$^2$ = 69.63, df = 10 (P < 0.00001); I$^2$ = 86%
Test for overall effect: Z = 1.55 (P = 0.12)

Risk Ratio
M-H, Random, 95% CI

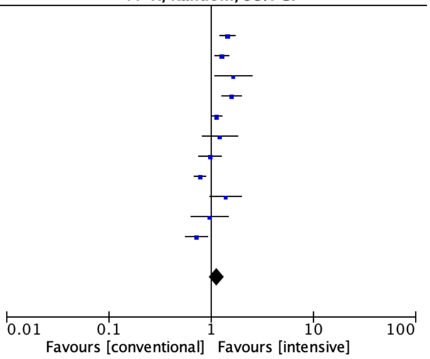


#### Leave out Anadani et al 2019

| | Intensive Management | | Conventional Management | | | Risk Ratio | |
|---|---|---|---|---|---|---|---|
| Study or Subgroup | Events | Total | Events | Total | Weight | M-H, Random, 95% CI | Year |
| Goyal et al 2017 | 7 | 10 | 67 | 130 | 6.8% | 1.36 [0.88, 2.11] | 2017 |
| Anadani et al 2019 | 485 | 942 | 92 | 259 | 0.0% | 1.45 [1.22, 1.73] | 2019 |
| Cernik D et al 2019 | 202 | 376 | 132 | 314 | 11.0% | 1.28 [1.09, 1.50] | 2019 |
| Chang et al 2019 | 32 | 48 | 17 | 42 | 7.1% | 1.65 [1.08, 2.50] | 2019 |
| Eva A. Mistry et al 2019 | 96 | 179 | 90 | 267 | 10.2% | 1.59 [1.28, 1.98] | 2019 |
| Anadani et al 2020 | 283 | 540 | 222 | 479 | 11.5% | 1.13 [1.00, 1.28] | 2020 |
| Han et al 2021 | 38 | 96 | 27 | 83 | 7.4% | 1.22 [0.82, 1.81] | 2021 |
| Mazighi et al 2021 | 67 | 152 | 69 | 153 | 9.7% | 0.98 [0.76, 1.26] | 2021 |
| Yang et al 2022 | 192 | 404 | 247 | 406 | 11.4% | 0.78 [0.69, 0.89] | 2022 |
| P. Upadhyaya et al 2023 | 28 | 44 | 27 | 59 | 8.0% | 1.39 [0.97, 1.99] | 2023 |
| Mistry et al 2023 | 17 | 37 | 36 | 76 | 7.0% | 0.97 [0.64, 1.48] | 2023 |
| Nam et al 2023 | 61 | 155 | 80 | 147 | 9.8% | 0.72 [0.57, 0.92] | 2023 |
| **Total (95% CI)** | | **2041** | | **2156** | **100.0%** | **1.13 [0.95, 1.34]** | |
| Total events | 1023 | | 1014 | | | | |

Heterogeneity: Tau$^2$ = 0.06; Chi$^2$ = 59.51, df = 10 (P < 0.00001); I$^2$ = 83%
Test for overall effect: Z = 1.39 (P = 0.17)

Risk Ratio
M-H, Random, 95% CI

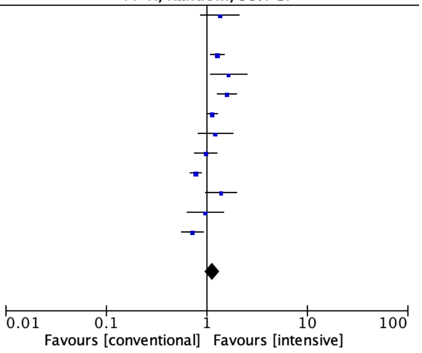


#### Leave out Cernik et al 2019

| | Intensive Management | | Conventional Management | | | Risk Ratio | |
|---|---|---|---|---|---|---|---|
| Study or Subgroup | Events | Total | Events | Total | Weight | M-H, Random, 95% CI | Year |
| Goyal et al 2017 | 7 | 10 | 67 | 130 | 7.0% | 1.36 [0.88, 2.11] | 2017 |
| Anadani et al 2019 | 485 | 942 | 92 | 259 | 10.7% | 1.45 [1.22, 1.73] | 2019 |
| Cernik D et al 2019 | 202 | 376 | 132 | 314 | 0.0% | 1.28 [1.09, 1.50] | 2019 |
| Chang et al 2019 | 32 | 48 | 17 | 42 | 7.3% | 1.65 [1.08, 2.50] | 2019 |
| Eva A. Mistry et al 2019 | 96 | 179 | 90 | 267 | 10.1% | 1.59 [1.28, 1.98] | 2019 |
| Anadani et al 2020 | 283 | 540 | 222 | 479 | 11.2% | 1.13 [1.00, 1.28] | 2020 |
| Han et al 2021 | 38 | 96 | 27 | 83 | 7.6% | 1.22 [0.82, 1.81] | 2021 |
| Mazighi et al 2021 | 67 | 152 | 69 | 153 | 9.7% | 0.98 [0.76, 1.26] | 2021 |
| Yang et al 2022 | 192 | 404 | 247 | 406 | 11.2% | 0.78 [0.69, 0.89] | 2022 |
| P. Upadhyaya et al 2023 | 28 | 44 | 27 | 59 | 8.2% | 1.39 [0.97, 1.99] | 2023 |
| Mistry et al 2023 | 17 | 37 | 36 | 76 | 7.3% | 0.97 [0.64, 1.48] | 2023 |
| Nam et al 2023 | 61 | 155 | 80 | 147 | 9.8% | 0.72 [0.57, 0.92] | 2023 |
| **Total (95% CI)** | | **2607** | | **2101** | **100.0%** | **1.15 [0.95, 1.38]** | |
| Total events | 1306 | | 974 | | | | |

Heterogeneity: Tau$^2$ = 0.07; Chi$^2$ = 66.62, df = 10 (P < 0.00001); I$^2$ = 85%
Test for overall effect: Z = 1.46 (P = 0.14)

Risk Ratio
M-H, Random, 95% CI

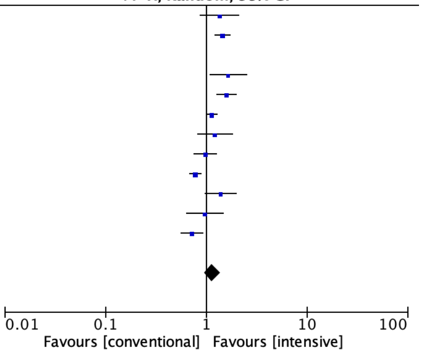

#### Leave out Chang et al 2019

| Study or Subgroup | Intensive Management Events | Intensive Management Total | Conventional Management Events | Conventional Management Total | Weight | Risk Ratio M-H, Random, 95% CI | Year |
|---|---|---|---|---|---|---|---|
| Goyal et al 2017 | 7 | 10 | 67 | 130 | 6.6% | 1.36 [0.88, 2.11] | 2017 |
| Anadani et al 2019 | 485 | 942 | 92 | 259 | 10.4% | 1.45 [1.22, 1.73] | 2019 |
| Cernik D et al 2019 | 202 | 376 | 132 | 314 | 10.6% | 1.28 [1.09, 1.50] | 2019 |
| Chang et al 2019 | 32 | 48 | 17 | 42 | 0.0% | 1.65 [1.08, 2.50] | 2019 |
| Eva A. Mistry et al 2019 | 96 | 179 | 90 | 267 | 9.9% | 1.59 [1.28, 1.98] | 2019 |
| Anadani et al 2020 | 283 | 540 | 222 | 479 | 11.0% | 1.13 [1.00, 1.28] | 2020 |
| Han et al 2021 | 38 | 96 | 27 | 83 | 7.2% | 1.22 [0.82, 1.81] | 2021 |
| Mazighi et al 2021 | 67 | 152 | 69 | 153 | 9.4% | 0.98 [0.76, 1.26] | 2021 |
| Yang et al 2022 | 192 | 404 | 247 | 406 | 11.0% | 0.78 [0.69, 0.89] | 2022 |
| P. Upadhyaya et al 2023 | 28 | 44 | 27 | 59 | 7.7% | 1.39 [0.97, 1.99] | 2023 |
| Mistry et al 2023 | 17 | 37 | 36 | 76 | 6.8% | 0.97 [0.64, 1.48] | 2023 |
| Nam et al 2023 | 61 | 155 | 80 | 147 | 9.4% | 0.72 [0.57, 0.92] | 2023 |
| **Total (95% CI)** | | **2935** | | **2373** | **100.0%** | **1.13 [0.95, 1.34]** | |
| Total events | 1476 | | 1089 | | | | |

Heterogeneity: Tau² = 0.06; Chi² = 66.82, df = 10 (P < 0.00001); I² = 85%
Test for overall effect: Z = 1.41 (P = 0.16)

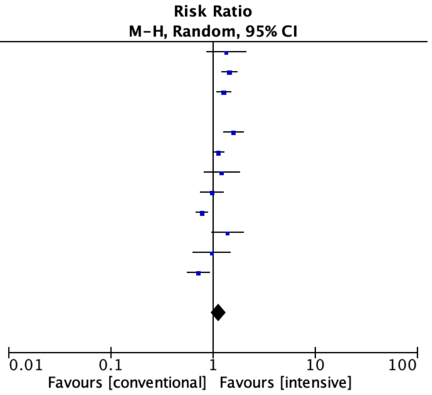


#### Leave out Mistry et al 2019

| Study or Subgroup | Intensive Management Events | Intensive Management Total | Conventional Management Events | Conventional Management Total | Weight | Risk Ratio M-H, Random, 95% CI | Year |
|---|---|---|---|---|---|---|---|
| Goyal et al 2017 | 7 | 10 | 67 | 130 | 6.6% | 1.36 [0.88, 2.11] | 2017 |
| Anadani et al 2019 | 485 | 942 | 92 | 259 | 10.9% | 1.45 [1.22, 1.73] | 2019 |
| Cernik D et al 2019 | 202 | 376 | 132 | 314 | 11.1% | 1.28 [1.09, 1.50] | 2019 |
| Chang et al 2019 | 32 | 48 | 17 | 42 | 6.9% | 1.65 [1.08, 2.50] | 2019 |
| Eva A. Mistry et al 2019 | 96 | 179 | 90 | 267 | 0.0% | 1.59 [1.28, 1.98] | 2019 |
| Anadani et al 2020 | 283 | 540 | 222 | 479 | 11.5% | 1.13 [1.00, 1.28] | 2020 |
| Han et al 2021 | 38 | 96 | 27 | 83 | 7.3% | 1.22 [0.82, 1.81] | 2021 |
| Mazighi et al 2021 | 67 | 152 | 69 | 153 | 9.7% | 0.98 [0.76, 1.26] | 2021 |
| Yang et al 2022 | 192 | 404 | 247 | 406 | 11.5% | 0.78 [0.69, 0.89] | 2022 |
| P. Upadhyaya et al 2023 | 28 | 44 | 27 | 59 | 7.9% | 1.39 [0.97, 1.99] | 2023 |
| Mistry et al 2023 | 17 | 37 | 36 | 76 | 6.9% | 0.97 [0.64, 1.48] | 2023 |
| Nam et al 2023 | 61 | 155 | 80 | 147 | 9.7% | 0.72 [0.57, 0.92] | 2023 |
| **Total (95% CI)** | | **2804** | | **2148** | **100.0%** | **1.12 [0.95, 1.32]** | |
| Total events | 1412 | | 1016 | | | | |

Heterogeneity: Tau² = 0.06; Chi² = 58.39, df = 10 (P < 0.00001); I² = 83%
Test for overall effect: Z = 1.33 (P = 0.18)

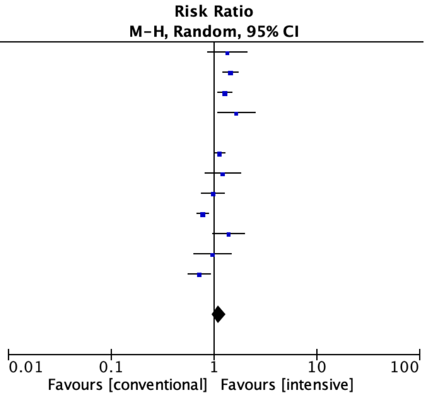


#### Leave out Anadani et al 2020

| Study or Subgroup | Intensive Management Events | Intensive Management Total | Conventional Management Events | Conventional Management Total | Weight | Risk Ratio M-H, Random, 95% CI | Year |
|---|---|---|---|---|---|---|---|
| Goyal et al 2017 | 7 | 10 | 67 | 130 | 7.3% | 1.36 [0.88, 2.11] | 2017 |
| Anadani et al 2019 | 485 | 942 | 92 | 259 | 10.5% | 1.45 [1.22, 1.73] | 2019 |
| Cernik D et al 2019 | 202 | 376 | 132 | 314 | 10.7% | 1.28 [1.09, 1.50] | 2019 |
| Chang et al 2019 | 32 | 48 | 17 | 42 | 7.5% | 1.65 [1.08, 2.50] | 2019 |
| Eva A. Mistry et al 2019 | 96 | 179 | 90 | 267 | 10.1% | 1.59 [1.28, 1.98] | 2019 |
| Anadani et al 2020 | 283 | 540 | 222 | 479 | 0.0% | 1.13 [1.00, 1.28] | 2020 |
| Han et al 2021 | 38 | 96 | 27 | 83 | 7.8% | 1.22 [0.82, 1.81] | 2021 |
| Mazighi et al 2021 | 67 | 152 | 69 | 153 | 9.7% | 0.98 [0.76, 1.26] | 2021 |
| Yang et al 2022 | 192 | 404 | 247 | 406 | 11.0% | 0.78 [0.69, 0.89] | 2022 |
| P. Upadhyaya et al 2023 | 28 | 44 | 27 | 59 | 8.3% | 1.39 [0.97, 1.99] | 2023 |
| Mistry et al 2023 | 17 | 37 | 36 | 76 | 7.5% | 0.97 [0.64, 1.48] | 2023 |
| Nam et al 2023 | 61 | 155 | 80 | 147 | 9.7% | 0.72 [0.57, 0.92] | 2023 |
| **Total (95% CI)** | | **2443** | | **1936** | **100.0%** | **1.16 [0.96, 1.41]** | |
| Total events | 1225 | | 884 | | | | |

Heterogeneity: Tau² = 0.09; Chi² = 70.32, df = 10 (P < 0.00001); I² = 86%
Test for overall effect: Z = 1.52 (P = 0.13)

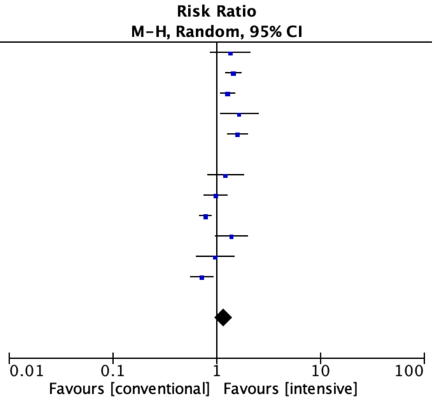


#### Leave out Han et al 2021

| Study or Subgroup | Intensive Management Events | Intensive Management Total | Conventional Management Events | Conventional Management Total | Weight | Risk Ratio M-H, Random, 95% CI | Year |
|---|---|---|---|---|---|---|---|
| Goyal et al 2017 | 7 | 10 | 67 | 130 | 6.7% | 1.36 [0.88, 2.11] | 2017 |
| Anadani et al 2019 | 485 | 942 | 92 | 259 | 10.4% | 1.45 [1.22, 1.73] | 2019 |
| Cernik D et al 2019 | 202 | 376 | 132 | 314 | 10.6% | 1.28 [1.09, 1.50] | 2019 |
| Chang et al 2019 | 32 | 48 | 17 | 42 | 7.0% | 1.65 [1.08, 2.50] | 2019 |
| Eva A. Mistry et al 2019 | 96 | 179 | 90 | 267 | 9.9% | 1.59 [1.28, 1.98] | 2019 |
| Anadani et al 2020 | 283 | 540 | 222 | 479 | 11.0% | 1.13 [1.00, 1.28] | 2020 |
| Han et al 2021 | 38 | 96 | 27 | 83 | 0.0% | 1.22 [0.82, 1.81] | 2021 |
| Mazighi et al 2021 | 67 | 152 | 69 | 153 | 9.4% | 0.98 [0.76, 1.26] | 2021 |
| Yang et al 2022 | 192 | 404 | 247 | 406 | 10.9% | 0.78 [0.69, 0.89] | 2022 |
| P. Upadhyaya et al 2023 | 28 | 44 | 27 | 59 | 7.8% | 1.39 [0.97, 1.99] | 2023 |
| Mistry et al 2023 | 17 | 37 | 36 | 76 | 6.9% | 0.97 [0.64, 1.48] | 2023 |
| Nam et al 2023 | 61 | 155 | 80 | 147 | 9.5% | 0.72 [0.57, 0.92] | 2023 |
| **Total (95% CI)** | | **2887** | | **2332** | **100.0%** | **1.15 [0.97, 1.37]** | |
| Total events | 1470 | | 1079 | | | | |

Heterogeneity: Tau² = 0.07; Chi² = 70.20, df = 10 (P < 0.00001); I² = 86%
Test for overall effect: Z = 1.61 (P = 0.11)

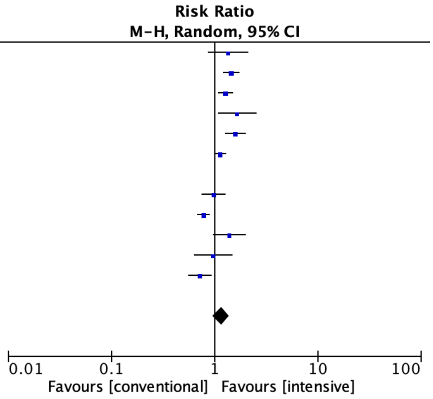

### Leave out Mazighi et al

| Study or Subgroup | Intensive Management Events | Intensive Management Total | Conventional Management Events | Conventional Management Total | Weight | Risk Ratio M-H, Random, 95% CI | Year |
|---|---|---|---|---|---|---|---|
| Goyal et al 2017 | 7 | 10 | 67 | 130 | 6.9% | 1.36 [0.88, 2.11] | 2017 |
| Anadani et al 2019 | 485 | 942 | 92 | 259 | 10.6% | 1.45 [1.22, 1.73] | 2019 |
| Cernik D et al 2019 | 202 | 376 | 132 | 314 | 10.8% | 1.28 [1.09, 1.50] | 2019 |
| Chang et al 2019 | 32 | 48 | 17 | 42 | 7.2% | 1.65 [1.08, 2.50] | 2019 |
| Eva A. Mistry et al 2019 | 96 | 179 | 90 | 267 | 10.1% | 1.59 [1.28, 1.98] | 2019 |
| Anadani et al 2020 | 283 | 540 | 222 | 479 | 11.2% | 1.13 [1.00, 1.28] | 2020 |
| Han et al 2021 | 38 | 96 | 27 | 83 | 7.5% | 1.22 [0.82, 1.81] | 2021 |
| Mazighi et al 2021 | 67 | 152 | 69 | 153 | 0.0% | 0.98 [0.76, 1.26] | 2021 |
| Yang et al 2022 | 192 | 404 | 247 | 406 | 11.1% | 0.78 [0.69, 0.89] | 2022 |
| P. Upadhyaya et al 2023 | 28 | 44 | 27 | 59 | 8.0% | 1.39 [0.97, 1.99] | 2023 |
| Mistry et al 2023 | 17 | 37 | 36 | 76 | 7.1% | 0.97 [0.64, 1.48] | 2023 |
| Nam et al 2023 | 61 | 155 | 80 | 147 | 9.7% | 0.72 [0.57, 0.92] | 2023 |
| **Total (95% CI)** | | **2831** | | **2262** | **100.0%** | **1.18 [0.99, 1.41]** | |
| Total events | 1441 | | 1037 | | | | |

Heterogeneity: $Tau^2 = 0.07$; $Chi^2 = 69.57$, df = 10 (P < 0.00001); $I^2 = 86\%$
Test for overall effect: Z = 1.80 (P = 0.07)

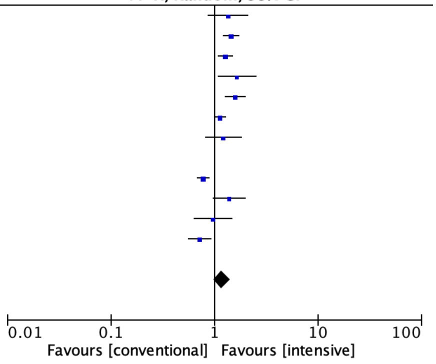


### Leave out Yang et al 2022*

| Study or Subgroup | Intensive Management Events | Intensive Management Total | Conventional Management Events | Conventional Management Total | Weight | Risk Ratio M-H, Random, 95% CI | Year |
|---|---|---|---|---|---|---|---|
| Goyal et al 2017 | 7 | 10 | 67 | 130 | 6.0% | 1.36 [0.88, 2.11] | 2017 |
| Anadani et al 2019 | 485 | 942 | 92 | 259 | 11.7% | 1.45 [1.22, 1.73] | 2019 |
| Cernik D et al 2019 | 202 | 376 | 132 | 314 | 12.1% | 1.28 [1.09, 1.50] | 2019 |
| Chang et al 2019 | 32 | 48 | 17 | 42 | 6.3% | 1.65 [1.08, 2.50] | 2019 |
| Eva A. Mistry et al 2019 | 96 | 179 | 90 | 267 | 10.7% | 1.59 [1.28, 1.98] | 2019 |
| Anadani et al 2020 | 283 | 540 | 222 | 479 | 12.9% | 1.13 [1.00, 1.28] | 2020 |
| Han et al 2021 | 38 | 96 | 27 | 83 | 6.7% | 1.22 [0.82, 1.81] | 2021 |
| Mazighi et al 2021 | 67 | 152 | 69 | 153 | 9.9% | 0.98 [0.76, 1.26] | 2021 |
| Yang et al 2022 | 192 | 404 | 247 | 406 | 0.0% | 0.78 [0.69, 0.89] | 2022 |
| P. Upadhyaya et al 2023 | 28 | 44 | 27 | 59 | 7.5% | 1.39 [0.97, 1.99] | 2023 |
| Mistry et al 2023 | 17 | 37 | 36 | 76 | 6.3% | 0.97 [0.64, 1.48] | 2023 |
| Nam et al 2023 | 61 | 155 | 80 | 147 | 10.0% | 0.72 [0.57, 0.92] | 2023 |
| **Total (95% CI)** | | **2579** | | **2009** | **100.0%** | **1.21 [1.05, 1.39]** | |
| Total events | 1316 | | 859 | | | | |

Heterogeneity: $Tau^2 = 0.04$; $Chi^2 = 35.55$, df = 10 (P = 0.0001); $I^2 = 72\%$
Test for overall effect: Z = 2.64 (P = 0.008)

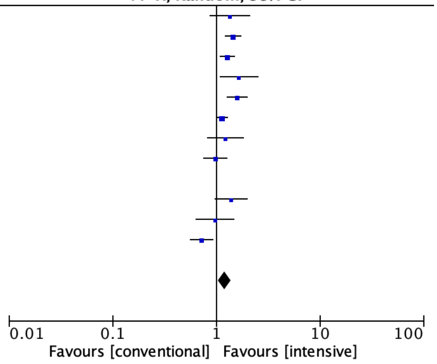


### Leave out Nam et al 2023*

| Study or Subgroup | Intensive Management Events | Intensive Management Total | Conventional Management Events | Conventional Management Total | Weight | Risk Ratio M-H, Random, 95% CI | Year |
|---|---|---|---|---|---|---|---|
| Goyal et al 2017 | 7 | 10 | 67 | 130 | 6.6% | 1.36 [0.88, 2.11] | 2017 |
| Anadani et al 2019 | 485 | 942 | 92 | 259 | 10.8% | 1.45 [1.22, 1.73] | 2019 |
| Cernik D et al 2019 | 202 | 376 | 132 | 314 | 11.1% | 1.28 [1.09, 1.50] | 2019 |
| Chang et al 2019 | 32 | 48 | 17 | 42 | 6.9% | 1.65 [1.08, 2.50] | 2019 |
| Eva A. Mistry et al 2019 | 96 | 179 | 90 | 267 | 10.2% | 1.59 [1.28, 1.98] | 2019 |
| Anadani et al 2020 | 283 | 540 | 222 | 479 | 11.5% | 1.13 [1.00, 1.28] | 2020 |
| Han et al 2021 | 38 | 96 | 27 | 83 | 7.2% | 1.22 [0.82, 1.81] | 2021 |
| Mazighi et al 2021 | 67 | 152 | 69 | 153 | 9.6% | 0.98 [0.76, 1.26] | 2021 |
| Yang et al 2022 | 192 | 404 | 247 | 406 | 11.5% | 0.78 [0.69, 0.89] | 2022 |
| P. Upadhyaya et al 2023 | 28 | 44 | 27 | 59 | 7.8% | 1.39 [0.97, 1.99] | 2023 |
| Mistry et al 2023 | 17 | 37 | 36 | 76 | 6.8% | 0.97 [0.64, 1.48] | 2023 |
| Nam et al 2023 | 61 | 155 | 80 | 147 | 0.0% | 0.72 [0.57, 0.92] | 2023 |
| **Total (95% CI)** | | **2828** | | **2268** | **100.0%** | **1.21 [1.03, 1.43]** | |
| Total events | 1447 | | 1026 | | | | |

Heterogeneity: $Tau^2 = 0.06$; $Chi^2 = 58.52$, df = 10 (P < 0.00001); $I^2 = 83\%$
Test for overall effect: Z = 2.27 (P = 0.02)

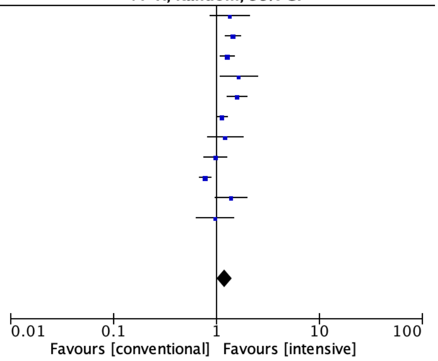

### Leave out Upadhyaya et al 2023

| Study or Subgroup | Intensive Management Events | Intensive Management Total | Conventional Management Events | Conventional Management Total | Weight | Risk Ratio M-H, Random, 95% CI | Year |
|---|---|---|---|---|---|---|---|
| Goyal et al 2017 | 7 | 10 | 67 | 130 | 6.7% | 1.36 [0.88, 2.11] | 2017 |
| Anadani et al 2019 | 485 | 942 | 92 | 259 | 10.5% | 1.45 [1.22, 1.73] | 2019 |
| Cernik D et al 2019 | 202 | 376 | 132 | 314 | 10.7% | 1.28 [1.09, 1.50] | 2019 |
| Chang et al 2019 | 32 | 48 | 17 | 42 | 7.0% | 1.65 [1.08, 2.50] | 2019 |
| Eva A. Mistry et al 2019 | 96 | 179 | 90 | 267 | 9.9% | 1.59 [1.28, 1.98] | 2019 |
| Anadani et al 2020 | 283 | 540 | 222 | 479 | 11.1% | 1.13 [1.00, 1.28] | 2020 |
| Han et al 2021 | 38 | 96 | 27 | 83 | 7.3% | 1.22 [0.82, 1.81] | 2021 |
| Mazighi et al 2021 | 67 | 152 | 69 | 153 | 9.4% | 0.98 [0.76, 1.26] | 2021 |
| Yang et al 2022 | 192 | 404 | 247 | 406 | 11.0% | 0.78 [0.69, 0.89] | 2022 |
| P. Upadhyaya et al 2023 | 28 | 44 | 27 | 59 | 0.0% | 1.39 [0.97, 1.99] | 2023 |
| Mistry et al 2023 | 17 | 37 | 36 | 76 | 6.9% | 0.97 [0.64, 1.48] | 2023 |
| Nam et al 2023 | 61 | 155 | 80 | 147 | 9.5% | 0.72 [0.57, 0.92] | 2023 |
| **Total (95% CI)** | | **2939** | | **2356** | **100.0%** | **1.14 [0.96, 1.36]** | |
| Total events | 1480 | | 1079 | | | | |

Heterogeneity: $Tau^2 = 0.07$; $Chi^2 = 68.81$, df = 10 ($P < 0.00001$); $I^2 = 85\%$
Test for overall effect: Z = 1.49 (P = 0.13)

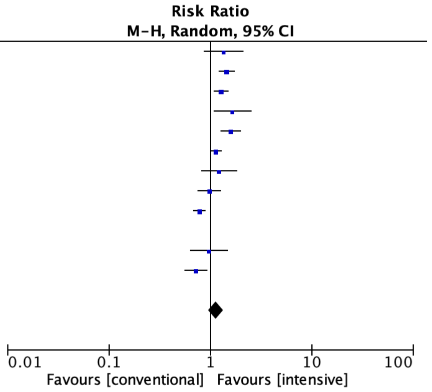


### Leave out Mistry et al 2023

| Study or Subgroup | Intensive Management Events | Intensive Management Total | Conventional Management Events | Conventional Management Total | Weight | Risk Ratio M-H, Random, 95% CI | Year |
|---|---|---|---|---|---|---|---|
| Goyal et al 2017 | 7 | 10 | 67 | 130 | 6.7% | 1.36 [0.88, 2.11] | 2017 |
| Anadani et al 2019 | 485 | 942 | 92 | 259 | 10.4% | 1.45 [1.22, 1.73] | 2019 |
| Cernik D et al 2019 | 202 | 376 | 132 | 314 | 10.6% | 1.28 [1.09, 1.50] | 2019 |
| Chang et al 2019 | 32 | 48 | 17 | 42 | 6.9% | 1.65 [1.08, 2.50] | 2019 |
| Eva A. Mistry et al 2019 | 96 | 179 | 90 | 267 | 9.8% | 1.59 [1.28, 1.98] | 2019 |
| Anadani et al 2020 | 283 | 540 | 222 | 479 | 10.9% | 1.13 [1.00, 1.28] | 2020 |
| Han et al 2021 | 38 | 96 | 27 | 83 | 7.2% | 1.22 [0.82, 1.81] | 2021 |
| Mazighi et al 2021 | 67 | 152 | 69 | 153 | 9.3% | 0.98 [0.76, 1.26] | 2021 |
| Yang et al 2022 | 192 | 404 | 247 | 406 | 10.9% | 0.78 [0.69, 0.89] | 2022 |
| P. Upadhyaya et al 2023 | 28 | 44 | 27 | 59 | 7.8% | 1.39 [0.97, 1.99] | 2023 |
| Mistry et al 2023 | 17 | 37 | 36 | 76 | 0.0% | 0.97 [0.64, 1.48] | 2023 |
| Nam et al 2023 | 61 | 155 | 80 | 147 | 9.4% | 0.72 [0.57, 0.92] | 2023 |
| **Total (95% CI)** | | **2946** | | **2339** | **100.0%** | **1.17 [0.99, 1.39]** | |
| Total events | 1491 | | 1070 | | | | |

Heterogeneity: $Tau^2 = 0.07$; $Chi^2 = 70.12$, df = 10 ($P < 0.00001$); $I^2 = 86\%$
Test for overall effect: Z = 1.80 (P = 0.07)

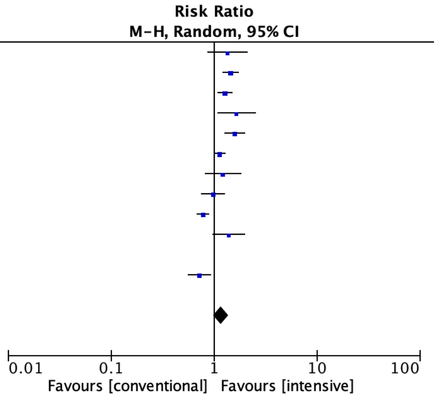


### Leave out one study analysis for primary outcome of observational studies

### Leave out Goyal et al 2017

| Study or Subgroup | Intensive Control Events | Intensive Control Total | Conventional Control Events | Conventional Control Total | Weight | Risk Ratio M-H, Random, 95% CI | Year |
|---|---|---|---|---|---|---|---|
| Goyal et al 2017 | 7 | 10 | 67 | 130 | 0.0% | 1.36 [0.88, 2.11] | 2017 |
| Eva A. Mistry et al 2019 | 96 | 179 | 90 | 267 | 15.4% | 1.59 [1.28, 1.98] | 2019 |
| Anadani et al 2019 | 485 | 942 | 92 | 259 | 19.0% | 1.45 [1.22, 1.73] | 2019 |
| Cernik D et al 2019 | 202 | 376 | 132 | 314 | 20.5% | 1.28 [1.09, 1.50] | 2019 |
| Chang et al 2019 | 32 | 48 | 17 | 42 | 6.1% | 1.65 [1.08, 2.50] | 2019 |
| Anadani et al 2020 | 283 | 540 | 222 | 479 | 24.3% | 1.13 [1.00, 1.28] | 2020 |
| Han et al 2021 | 38 | 96 | 27 | 83 | 6.7% | 1.22 [0.82, 1.81] | 2021 |
| P. Upadhyaya et al 2023 | 28 | 44 | 27 | 59 | 7.9% | 1.39 [0.97, 1.99] | 2023 |
| **Total (95% CI)** | | **2225** | | **1503** | **100.0%** | **1.34 [1.19, 1.50]** | |
| Total events | 1164 | | 607 | | | | |

Heterogeneity: $Tau^2 = 0.01$; $Chi^2 = 11.13$, df = 6 (P = 0.08); $I^2 = 46\%$
Test for overall effect: Z = 5.01 ($P < 0.00001$)

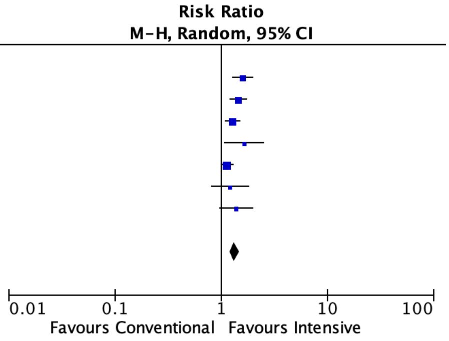

#### Leave out Anadani et al 2019

| Study or Subgroup | Intensive Control Events | Intensive Control Total | Conventional Control Events | Conventional Control Total | Weight | Risk Ratio M-H, Random, 95% CI | Year |
|---|---|---|---|---|---|---|---|
| Goyal et al 2017 | 7 | 10 | 67 | 130 | 6.1% | 1.36 [0.88, 2.11] | 2017 |
| Eva A. Mistry et al 2019 | 96 | 179 | 90 | 267 | 17.6% | 1.59 [1.28, 1.98] | 2019 |
| Anadani et al 2019 | 485 | 942 | 92 | 259 | 0.0% | 1.45 [1.22, 1.73] | 2019 |
| Cernik D et al 2019 | 202 | 376 | 132 | 314 | 24.2% | 1.28 [1.09, 1.50] | 2019 |
| Chang et al 2019 | 32 | 48 | 17 | 42 | 6.7% | 1.65 [1.08, 2.50] | 2019 |
| Anadani et al 2020 | 283 | 540 | 222 | 479 | 29.4% | 1.13 [1.00, 1.28] | 2020 |
| Han et al 2021 | 38 | 96 | 27 | 83 | 7.3% | 1.22 [0.82, 1.81] | 2021 |
| P. Upadhyaya et al 2023 | 28 | 44 | 27 | 59 | 8.7% | 1.39 [0.97, 1.99] | 2023 |
| **Total (95% CI)** | | **1293** | | **1374** | **100.0%** | **1.31 [1.17, 1.48]** | |
| Total events | 686 | | 582 | | | | |

Heterogeneity: $Tau^2 = 0.01$; $Chi^2 = 9.33$, df = 6 (P = 0.16); $I^2 = 36\%$
Test for overall effect: Z = 4.56 (P < 0.00001)

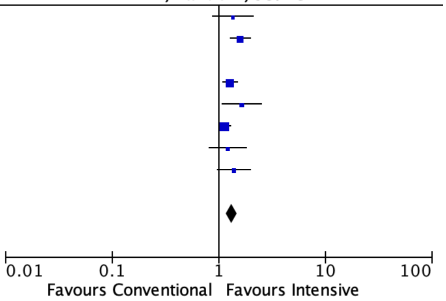


#### Leave out Cernik et al 2019

| Study or Subgroup | Intensive Control Events | Intensive Control Total | Conventional Control Events | Conventional Control Total | Weight | Risk Ratio M-H, Random, 95% CI | Year |
|---|---|---|---|---|---|---|---|
| Goyal et al 2017 | 7 | 10 | 67 | 130 | 7.2% | 1.36 [0.88, 2.11] | 2017 |
| Eva A. Mistry et al 2019 | 96 | 179 | 90 | 267 | 18.1% | 1.59 [1.28, 1.98] | 2019 |
| Anadani et al 2019 | 485 | 942 | 92 | 259 | 21.7% | 1.45 [1.22, 1.73] | 2019 |
| Cernik D et al 2019 | 202 | 376 | 132 | 314 | 0.0% | 1.28 [1.09, 1.50] | 2019 |
| Chang et al 2019 | 32 | 48 | 17 | 42 | 7.8% | 1.65 [1.08, 2.50] | 2019 |
| Anadani et al 2020 | 283 | 540 | 222 | 479 | 26.7% | 1.13 [1.00, 1.28] | 2020 |
| Han et al 2021 | 38 | 96 | 27 | 83 | 8.5% | 1.22 [0.82, 1.81] | 2021 |
| P. Upadhyaya et al 2023 | 28 | 44 | 27 | 59 | 9.9% | 1.39 [0.97, 1.99] | 2023 |
| **Total (95% CI)** | | **1859** | | **1319** | **100.0%** | **1.36 [1.19, 1.55]** | |
| Total events | 969 | | 542 | | | | |

Heterogeneity: $Tau^2 = 0.01$; $Chi^2 = 11.11$, df = 6 (P = 0.08); $I^2 = 46\%$
Test for overall effect: Z = 4.56 (P < 0.00001)

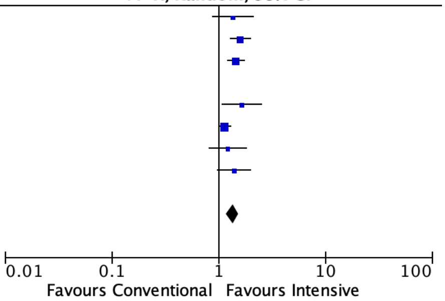


#### Leave out Chang et al 2019

| Study or Subgroup | Intensive Control Events | Intensive Control Total | Conventional Control Events | Conventional Control Total | Weight | Risk Ratio M-H, Random, 95% CI | Year |
|---|---|---|---|---|---|---|---|
| Goyal et al 2017 | 7 | 10 | 67 | 130 | 5.2% | 1.36 [0.88, 2.11] | 2017 |
| Eva A. Mistry et al 2019 | 96 | 179 | 90 | 267 | 15.2% | 1.59 [1.28, 1.98] | 2019 |
| Anadani et al 2019 | 485 | 942 | 92 | 259 | 19.3% | 1.45 [1.22, 1.73] | 2019 |
| Cernik D et al 2019 | 202 | 376 | 132 | 314 | 21.1% | 1.28 [1.09, 1.50] | 2019 |
| Chang et al 2019 | 32 | 48 | 17 | 42 | 0.0% | 1.65 [1.08, 2.50] | 2019 |
| Anadani et al 2020 | 283 | 540 | 222 | 479 | 25.7% | 1.13 [1.00, 1.28] | 2020 |
| Han et al 2021 | 38 | 96 | 27 | 83 | 6.2% | 1.22 [0.82, 1.81] | 2021 |
| P. Upadhyaya et al 2023 | 28 | 44 | 27 | 59 | 7.4% | 1.39 [0.97, 1.99] | 2023 |
| **Total (95% CI)** | | **2187** | | **1591** | **100.0%** | **1.32 [1.19, 1.47]** | |
| Total events | 1139 | | 657 | | | | |

Heterogeneity: $Tau^2 = 0.01$; $Chi^2 = 9.90$, df = 6 (P = 0.13); $I^2 = 39\%$
Test for overall effect: Z = 5.08 (P < 0.00001)

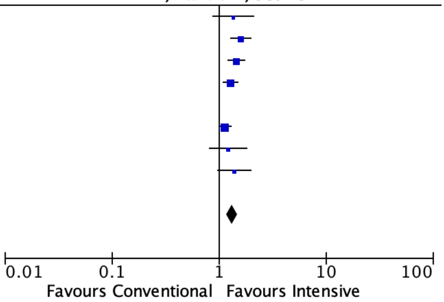


#### Leave out Mistry et al 2019

| Study or Subgroup | Intensive Control Events | Intensive Control Total | Conventional Control Events | Conventional Control Total | Weight | Risk Ratio M-H, Random, 95% CI | Year |
|---|---|---|---|---|---|---|---|
| Goyal et al 2017 | 7 | 10 | 67 | 130 | 4.4% | 1.36 [0.88, 2.11] | 2017 |
| Eva A. Mistry et al 2019 | 96 | 179 | 90 | 267 | 0.0% | 1.59 [1.28, 1.98] | 2019 |
| Anadani et al 2019 | 485 | 942 | 92 | 259 | 21.4% | 1.45 [1.22, 1.73] | 2019 |
| Cernik D et al 2019 | 202 | 376 | 132 | 314 | 24.3% | 1.28 [1.09, 1.50] | 2019 |
| Chang et al 2019 | 32 | 48 | 17 | 42 | 4.9% | 1.65 [1.08, 2.50] | 2019 |
| Anadani et al 2020 | 283 | 540 | 222 | 479 | 33.1% | 1.13 [1.00, 1.28] | 2020 |
| Han et al 2021 | 38 | 96 | 27 | 83 | 5.4% | 1.22 [0.82, 1.81] | 2021 |
| P. Upadhyaya et al 2023 | 28 | 44 | 27 | 59 | 6.5% | 1.39 [0.97, 1.99] | 2023 |
| **Total (95% CI)** | | **2056** | | **1366** | **100.0%** | **1.28 [1.17, 1.41]** | |
| Total events | 1075 | | 584 | | | | |

Heterogeneity: $Tau^2 = 0.00$; $Chi^2 = 7.38$, df = 6 (P = 0.29); $I^2 = 19\%$
Test for overall effect: Z = 5.14 (P < 0.00001)

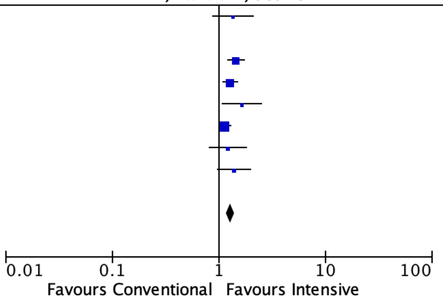

#### Leave out Anadani et 2020

| Study or Subgroup | Intensive Control Events | Intensive Control Total | Conventional Control Events | Conventional Control Total | Weight | Risk Ratio M-H, Random, 95% CI | Year |
|---|---|---|---|---|---|---|---|
| Goyal et al 2017 | 7 | 10 | 67 | 130 | 4.4% | 1.36 [0.88, 2.11] | 2017 |
| Eva A. Mistry et al 2019 | 96 | 179 | 90 | 267 | 18.1% | 1.59 [1.28, 1.98] | 2019 |
| Anadani et al 2019 | 485 | 942 | 92 | 259 | 27.6% | 1.45 [1.22, 1.73] | 2019 |
| Cernik D et al 2019 | 202 | 376 | 132 | 314 | 33.0% | 1.28 [1.09, 1.50] | 2019 |
| Chang et al 2019 | 32 | 48 | 17 | 42 | 4.9% | 1.65 [1.08, 2.50] | 2019 |
| Anadani et al 2020 | 283 | 540 | 222 | 479 | 0.0% | 1.13 [1.00, 1.28] | 2020 |
| Han et al 2021 | 38 | 96 | 27 | 83 | 5.4% | 1.22 [0.82, 1.81] | 2021 |
| P. Upadhyaya et al 2023 | 28 | 44 | 27 | 59 | 6.7% | 1.39 [0.97, 1.99] | 2023 |
| **Total (95% CI)** | | **1695** | | **1154** | **100.0%** | **1.40 [1.28, 1.54]** | |
| Total events | 888 | | 452 | | | | |

Heterogeneity: $Tau^2 = 0.00$; $Chi^2 = 3.82$, df = 6 (P = 0.70); $I^2 = 0\%$

Test for overall effect: Z = 7.19 (P < 0.00001)

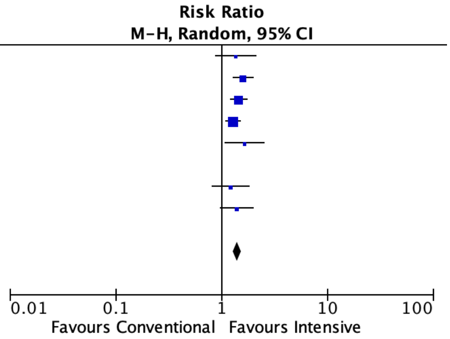


#### Leave out Han et al 2021

| Study or Subgroup | Intensive Control Events | Intensive Control Total | Conventional Control Events | Conventional Control Total | Weight | Risk Ratio M-H, Random, 95% CI | Year |
|---|---|---|---|---|---|---|---|
| Goyal et al 2017 | 7 | 10 | 67 | 130 | 5.7% | 1.36 [0.88, 2.11] | 2017 |
| Eva A. Mistry et al 2019 | 96 | 179 | 90 | 267 | 15.6% | 1.59 [1.28, 1.98] | 2019 |
| Anadani et al 2019 | 485 | 942 | 92 | 259 | 19.2% | 1.45 [1.22, 1.73] | 2019 |
| Cernik D et al 2019 | 202 | 376 | 132 | 314 | 20.7% | 1.28 [1.09, 1.50] | 2019 |
| Chang et al 2019 | 32 | 48 | 17 | 42 | 6.2% | 1.65 [1.08, 2.50] | 2019 |
| Anadani et al 2020 | 283 | 540 | 222 | 479 | 24.6% | 1.13 [1.00, 1.28] | 2020 |
| Han et al 2021 | 38 | 96 | 27 | 83 | 0.0% | 1.22 [0.82, 1.81] | 2021 |
| P. Upadhyaya et al 2023 | 28 | 44 | 27 | 59 | 8.0% | 1.39 [0.97, 1.99] | 2023 |
| **Total (95% CI)** | | **2139** | | **1550** | **100.0%** | **1.35 [1.20, 1.51]** | |
| Total events | 1133 | | 647 | | | | |

Heterogeneity: $Tau^2 = 0.01$; $Chi^2 = 11.06$, df = 6 (P = 0.09); $I^2 = 46\%$

Test for overall effect: Z = 5.12 (P < 0.00001)

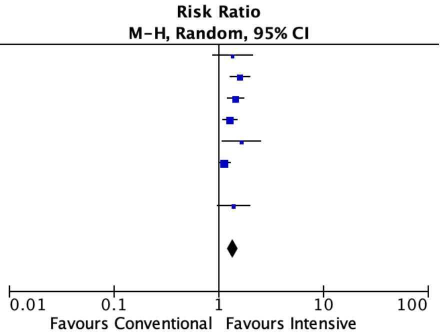


#### Leave out Upadhyaya et al 2023

| Study or Subgroup | Intensive Control Events | Intensive Control Total | Conventional Control Events | Conventional Control Total | Weight | Risk Ratio M-H, Random, 95% CI | Year |
|---|---|---|---|---|---|---|---|
| Goyal et al 2017 | 7 | 10 | 67 | 130 | 5.8% | 1.36 [0.88, 2.11] | 2017 |
| Eva A. Mistry et al 2019 | 96 | 179 | 90 | 267 | 15.8% | 1.59 [1.28, 1.98] | 2019 |
| Anadani et al 2019 | 485 | 942 | 92 | 259 | 19.4% | 1.45 [1.22, 1.73] | 2019 |
| Cernik D et al 2019 | 202 | 376 | 132 | 314 | 21.0% | 1.28 [1.09, 1.50] | 2019 |
| Chang et al 2019 | 32 | 48 | 17 | 42 | 6.3% | 1.65 [1.08, 2.50] | 2019 |
| Anadani et al 2020 | 283 | 540 | 222 | 479 | 24.9% | 1.13 [1.00, 1.28] | 2020 |
| Han et al 2021 | 38 | 96 | 27 | 83 | 6.9% | 1.22 [0.82, 1.81] | 2021 |
| P. Upadhyaya et al 2023 | 28 | 44 | 27 | 59 | 0.0% | 1.39 [0.97, 1.99] | 2023 |
| **Total (95% CI)** | | **2191** | | **1574** | **100.0%** | **1.34 [1.19, 1.50]** | |
| Total events | 1143 | | 647 | | | | |

Heterogeneity: $Tau^2 = 0.01$; $Chi^2 = 11.03$, df = 6 (P = 0.09); $I^2 = 46\%$

Test for overall effect: Z = 4.92 (P < 0.00001)

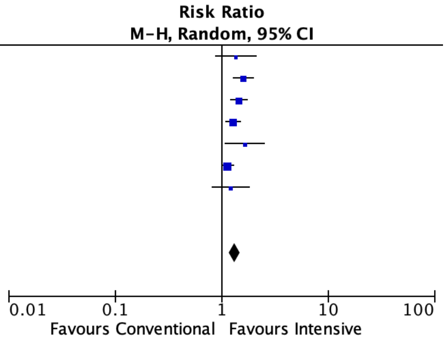

Leave out one study analysis for RCTs

#### Leave out Yang et al 2022*

| | Intensive Control | | Conventional Control | | | Risk Ratio | |
|---|---|---|---|---|---|---|---|
| Study or Subgroup | Events | Total | Events | Total | Weight | M-H, Random, 95% CI | Year |
| Mazighi et al 2021 | 67 | 152 | 69 | 153 | 39.7% | 0.98 [0.76, 1.26] | 2021 |
| Yang et al 2022 | 192 | 404 | 247 | 406 | 0.0% | 0.78 [0.69, 0.89] | 2022 |
| Nam et al 2023 | 61 | 155 | 80 | 147 | 40.6% | 0.72 [0.57, 0.92] | 2023 |
| Mistry et al 2023 | 17 | 37 | 36 | 76 | 19.8% | 0.97 [0.64, 1.48] | 2023 |
| **Total (95% CI)** | | **344** | | **376** | **100.0%** | **0.86 [0.70, 1.07]** | |
| Total events | 145 | | 185 | | | | |

Heterogeneity: Tau$^2$ = 0.01; Chi$^2$ = 3.23, df = 2 (P = 0.20); I$^2$ = 38%
Test for overall effect: Z = 1.34 (P = 0.18)

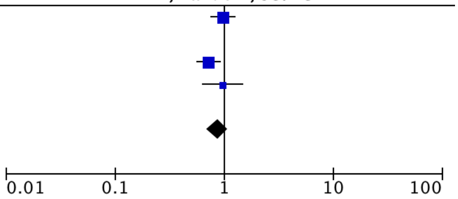


#### Leave out Nam et al 2023*

| | Intensive Control | | Conventional Control | | | Risk Ratio | |
|---|---|---|---|---|---|---|---|
| Study or Subgroup | Events | Total | Events | Total | Weight | M-H, Random, 95% CI | Year |
| Mazighi et al 2021 | 67 | 152 | 69 | 153 | 29.2% | 0.98 [0.76, 1.26] | 2021 |
| Yang et al 2022 | 192 | 404 | 247 | 406 | 57.8% | 0.78 [0.69, 0.89] | 2022 |
| Nam et al 2023 | 61 | 155 | 80 | 147 | 0.0% | 0.72 [0.57, 0.92] | 2023 |
| Mistry et al 2023 | 17 | 37 | 36 | 76 | 13.0% | 0.97 [0.64, 1.48] | 2023 |
| **Total (95% CI)** | | **593** | | **635** | **100.0%** | **0.86 [0.73, 1.01]** | |
| Total events | 276 | | 352 | | | | |

Heterogeneity: Tau$^2$ = 0.01; Chi$^2$ = 3.03, df = 2 (P = 0.22); I$^2$ = 34%
Test for overall effect: Z = 1.83 (P = 0.07)

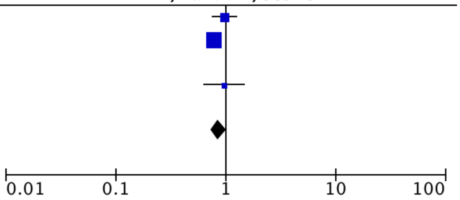


Leave out Mistry et al 2023

| | Intensive Control | | Conventional Control | | | Risk Ratio | |
|---|---|---|---|---|---|---|---|
| Study or Subgroup | Events | Total | Events | Total | Weight | M-H, Random, 95% CI | Year |
| Mazighi et al 2021 | 67 | 152 | 69 | 153 | 24.3% | 0.98 [0.76, 1.26] | 2021 |
| Yang et al 2022 | 192 | 404 | 247 | 406 | 50.6% | 0.78 [0.69, 0.89] | 2022 |
| Nam et al 2023 | 61 | 155 | 80 | 147 | 25.0% | 0.72 [0.57, 0.92] | 2023 |
| Mistry et al 2023 | 17 | 37 | 36 | 76 | 0.0% | 0.97 [0.64, 1.48] | 2023 |
| **Total (95% CI)** | | **711** | | **706** | **100.0%** | **0.81 [0.70, 0.94]** | |
| Total events | 320 | | 396 | | | | |

Heterogeneity: Tau$^2$ = 0.01; Chi$^2$ = 3.25, df = 2 (P = 0.20); I$^2$ = 38%
Test for overall effect: Z = 2.83 (P = 0.005)

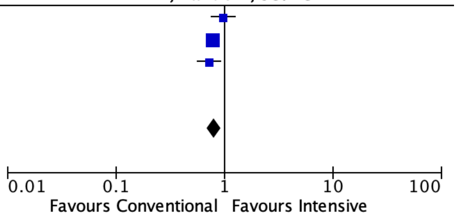


Leave out Mazighi et al 2021

| | Intensive Control | | Conventional Control | | | Risk Ratio | |
|---|---|---|---|---|---|---|---|
| Study or Subgroup | Events | Total | Events | Total | Weight | M-H, Random, 95% CI | Year |
| Mazighi et al 2021 | 67 | 152 | 69 | 153 | 0.0% | 0.98 [0.76, 1.26] | 2021 |
| Yang et al 2022 | 192 | 404 | 247 | 406 | 73.0% | 0.78 [0.69, 0.89] | 2022 |
| Nam et al 2023 | 61 | 155 | 80 | 147 | 20.2% | 0.72 [0.57, 0.92] | 2023 |
| Mistry et al 2023 | 17 | 37 | 36 | 76 | 6.8% | 0.97 [0.64, 1.48] | 2023 |
| **Total (95% CI)** | | **596** | | **629** | **100.0%** | **0.78 [0.70, 0.87]** | |
| Total events | 270 | | 363 | | | | |

Heterogeneity: Tau$^2$ = 0.00; Chi$^2$ = 1.39, df = 2 (P = 0.50); I$^2$ = 0%
Test for overall effect: Z = 4.41 (P < 0.0001)

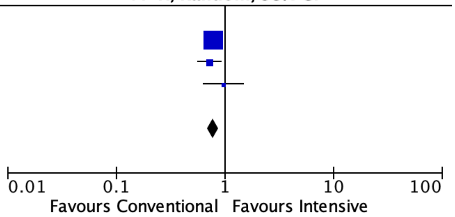

**GRADE Summary of findings:**

#### Intensive SBP Control compared to Conventional SBP Control for Patients with Acute Ischemic Stroke undergoing thrombectomy

**Patient or population:** Patients with Acute Ischemic Stroke undergoing thrombectomy
**Setting:** Acute ischemic Stroke undergoing thrombectomy
**Intervention:** Intensive SBP Control
**Comparison:** Conventional SBP Control

| Outcomes | Anticipated absolute effects* (95% CI) | | Relative effect (95% CI) | № of participants (studies) | Certainty of the evidence (GRADE) | Comments |
|---|---|---|---|---|---|---|
| | Risk with Conventional SBP Control | Risk with Intensive SBP Control | | | | |
| mRS 0-2 at 90 days | 413 per 1,000 | **549 per 1,000** (499 to 594) | **RR 1.33** (1.21 to 1.44) | 3868 (8 non-randomised studies) | ⨁⨁⨁◯ Moderate[a,b] | Intensive SBP Control probably results in an increased chances of mRS 0-2 at 90 days. |
| Death at 90 days | 230 per 1,000 | **165 per 1,000** (133 to 202) | **RR 0.72** (0.58 to 0.88) | 3868 (8 non-randomised studies) | ⨁⨁⨁◯ Moderate[a,b] | Intensive SBP Control likely reduces death at 90 days. |
| Symptomatic ICH | 75 per 1,000 | **47 per 1,000** (32 to 68) | **RR 0.62** (0.42 to 0.90) | 2642 (5 non-randomised studies) | ⨁⨁◯◯ Low[a,b,c] | Intensive SBP Control may reduce symptomatic ICH. |
| Decompressive hemicraniectomy | 68 per 1,000 | **26 per 1,000** (17 to 40) | **RR 0.39** (0.25 to 0.59) | 2323 (3 non-randomised studies) | ⨁⨁◯◯ Low[a,b,d] | Intensive SBP Control may reduce decompressive hemicraniectomy. |
| mRS 0-2 at 90 days (RCTs) | 552 per 1,000 | **459 per 1,000** (387 to 530) | **RR 0.83** (0.70 to 0.96) | 1530 (4 RCTs) | ⨁⨁⨁◯ Moderate[e] | Conventional SBP control results in improved chances of mRS 0-2 at 90 days |
| Death at 90 days | 140 per 1,000 | **159 per 1,000** (125 to 202) | **RR 1.13** (0.89 to 1.44) | 1534 (4 RCTs) | ⨁⨁⨁◯ Moderate[e] | No effect on mortality at 90 days between both arms. |
| Symptomatic ICH | 65 per 1,000 | **73 per 1,000** (51 to 105) | **RR 1.13** (0.78 to 1.62) | 1540 (4 RCTs) | ⨁⨁⨁◯ Moderate[e] | Neither Intensive nor conventional SBP Control result in any difference in symptomatic ICH rates. |

***The risk in the intervention group** (and its 95% confidence interval) is based on the assumed risk in the comparison group and the **relative effect** of the intervention (and its 95% CI).

**CI:** confidence interval; **RR:** risk ratio

**GRADE Working Group grades of evidence**
**High certainty:** we are very confident that the true effect lies close to that of the estimate of the effect.
**Moderate certainty:** we are moderately confident in the effect estimate: the true effect is likely to be close to the estimate of the effect, but there is a possibility that it is substantially different.
**Low certainty:** our confidence in the effect estimate is limited: the true effect may be substantially different from the estimate of the effect.
**Very low certainty:** we have very little confidence in the effect estimate: the true effect is likely to be substantially different from the estimate of effect.

#### Explanations

a. Inconsistencies in BP recordings and usage of Mean SBP for defining the intensive arm. Measurement and Observational Bias possible.
b. Cut offs used across different studies different
c. Low event rates in Two Studies-Goyal et al only 1/10 patients in the intensive arm and in P.Upadhyaya et al only 1/44 patients had symptomatic ICH
d. Low event rates and similar rates in P.Upadhyaya et al-1/44 in intensive arm and 1/59 in conventional arm
e. Deviations from intended BP Control arm-Domain 2

### GRADE EVIDENCE PROFILE

**Author(s): Baikuntha Panigrahi, Rohit Bhatia, Partha Haldar**
**Question:** Intensive SBP Control compared to Conventional SBP Control for Patients with Acute Ischemic Stroke undergoing thrombectomy
**Setting:** Acute ischemic Stroke undergoing thrombectomy
**Bibliography:**

| Certainty assessment | | | | | | | № of patients | | Effect | | Certainty | Importance |
|---|---|---|---|---|---|---|---|---|---|---|---|---|
| № of studies | Study design | Risk of bias | Inconsistency | Indirectness | Imprecision | Other considerations | Intensive SBP Control | Conventional SBP Control | Relative (95% CI) | Absolute (95% CI) | | |
| **mRS 0-2 at 90 days** | | | | | | | | | | | | |
| 8 | non-randomised studies | serious[a,b] | not serious | not serious | not serious | none | 1171/2235 (52.4%) | 674/1633 (41.3%) | **RR 1.33** (1.21 to 1.44) | **136 more per 1,000** (from 87 more to 182 more) | ⊕⊕⊕◯ Moderate | |
| **Death at 90 days** | | | | | | | | | | | | |
| 8 | non-randomised studies | serious[a,b] | not serious | not serious | not serious | none | 373/2235 (16.7%) | 375/1633 (23.0%) | **RR 0.72** (0.58 to 0.88) | **64 fewer per 1,000** (from 96 fewer to 28 fewer) | ⊕⊕⊕◯ Moderate | |
| **Symptomatic ICH** | | | | | | | | | | | | |
| 5 | non-randomised studies | serious[a,b] | not serious | not serious | serious[c] | none | 74/1632 (4.5%) | 76/1010 (7.5%) | **RR 0.62** (0.42 to 0.90) | **29 fewer per 1,000** (from 44 fewer to 8 fewer) | ⊕⊕◯◯ Low | |
| **Decompressive hemicraniectomy** | | | | | | | | | | | | |
| 3 | non-randomised studies | serious[a,b] | not serious | not serious | serious[d] | none | 43/1526 (2.8%) | 54/797 (6.8%) | **RR 0.39** (0.25 to 0.59) | **41 fewer per 1,000** (from 51 fewer to 28 fewer) | ⊕⊕◯◯ Low | |
| **mRS 0-2 at 90 days (RCTs)** | | | | | | | | | | | | |
| 4 | randomised trials | serious[e] | not serious | not serious | not serious | none | 337/748 (45.1%) | 432/782 (55.2%) | **RR 0.83** (0.70 to 0.96) | **94 fewer per 1,000** (from 166 fewer to 22 fewer) | ⊕⊕⊕◯ Moderate | |
| **Death at 90 days** | | | | | | | | | | | | |

| Certainty assessment | | | | | | | № of patients | | Effect | | Certainty | Importance |
|---|---|---|---|---|---|---|---|---|---|---|---|---|
| № of studies | Study design | Risk of bias | Inconsistency | Indirectness | Imprecision | Other considerations | Intensive SBP Control | Conventional SBP Control | Relative (95% CI) | Absolute (95% CI) | | |
| 4 | randomised trials | serious[e] | not serious | not serious | not serious | none | 114/750 (15.2%) | 110/784 (14.0%) | **RR 1.13** (0.89 to 1.44) | **18 more per 1,000** (from 15 fewer to 62 more) | ⊕⊕⊕◯ Moderate | |
| **Symptomatic ICH** | | | | | | | | | | | | |
| 4 | randomised trials | serious[e] | not serious | not serious | not serious | none | 56/753 (7.4%) | 51/787 (6.5%) | **RR 1.13** (0.78 to 1.62) | **8 more per 1,000** (from 14 fewer to 40 more) | ⊕⊕⊕◯ Moderate | |

**CI:** confidence interval; **RR:** risk ratio

#### Explanations

a. Inconsistencies in BP recordings and usage of Mean SBP for defining the intensive arm. Measurement and Observational Bias possible.
b. Cut offs used across different studies different
c. Low event rates in Two Studies-Goyal et al only 1/10 patients in the intensive arm and in P.Upadhyaya et al only 1/44 patients had symptomatic ICH
d. Low event rates and similar rates in P.Upadhyaya et al-1/44 in intensive arm and 1/59 in conventional arm
e. Deviations from intended BP Control arm-Domain 2
